# A review of the effectiveness and experiences of welfare advice services co-located in health settings: a critical narrative systematic review

**DOI:** 10.1101/2021.10.13.21264938

**Authors:** Sian Reece, Trevor A Sheldon, Josie Dickerson, Kate E Pickett

**Author notes:** CORRESPONDING AUTHOR Sian Reece, Hull York Medical School, Hull York Medical School, Heslington Road, York, YO105DD.

## Abstract

The links between financial insecurity and poor health and wellbeing are well established. Individuals experiencing financial insecurity are also more likely to face challenges in accessing the support services they need. There is evidence of unequal uptake of welfare support and benefits, particularly in some ethnic minority groups. The COVID-19 pandemic has further exacerbated financial insecurity for the most vulnerable and action is needed to improve the support provided for those affected during the recovery from the pandemic.

One approach to improving uptake of benefits has been to deliver welfare services within health settings. This has the potential to increase income and possibly improve health. We conducted systematic review with a critical narrative synthesis to assess the health, social and financial impacts of welfare advice services co-located in health settings and explore the facilitators and barriers to successful implementation of these services, in order to guide future policy and practice.

The review identified 14 studies published in the UK from 2010. The services provided generated on average £27 of social, economic and environmental return on investment per £1 invested. Individuals on average benefitted from an additional £2,757 household income per annum and cost savings for the NHS were demonstrated. The review demonstrated that improvements to health were made by addressing key social determinants of health, thereby reducing health inequalities. Co-located welfare services actively incorporated elements of proportionate universalism and targeted those, who due to predominately health needs, were most in need of this support. The nature of the welfare advice service, how it operates within a health setting, and how visible and accessible this service is to participants and professionals referring into the service, were seen as important facilitators. Co-production during service development and ongoing enhanced multi-disciplinary collaboration were also considered vital to the success of co-located services.

## INTRODUCTION

Early childhood deprivation is associated with significant negative physical, mental health and social outcomes that not only limit a child’s development in the short-term but have long lasting effects into adulthood (Marmot, Atkinson et al. 2010, Wickham, Anwar et al. 2016). In adulthood, links between financial insecurity, social deprivation and mental health are also well established (Marmot, Atkinson et al. 2010). Financial insecurity can precipitate and perpetuate mental health problems and has been found to be a predictor of chronic physical illness (Kahn and Pearlin 2006, Georgiades, Janszky et al. 2009, Advice Services Alliance 2015). Furthermore, individuals suffering with poor mental health associated with financial insecurity, exacerbated in recent years by austerity, are more likely to face challenges in accessing the advice and support needed to address these welfare issues (Jenkins, Bebbington et al. 2009, Fitch, Hamilton et al. 2011). The Covid-19 pandemic and other austerity measures have created and exacerbated financial insecurity for many families, further exacerbating existing inequalities (Dickerson, Kelly et al. 2021).

The adverse contribution of chronic financial insecurity to physical and mental health can be obviated if corrected early on (Kahn and Pearlin 2006). However, there is evidence of unequal access to benefits in some communities in the United Kingdom (UK), and this has been found to be particularly pronounced in some ethnic minority groups (Prady, Bloor et al. 2016).

Various schemes have been put in place to improve uptake of benefits by co-locating welfare rights advice services within health settings (Bateman 2008, Krska, Palmer et al. 2013, Woodhead, Khondoker et al. 2017). A systematic review of welfare rights advice delivered in health settings found that there was evidence that this approach resulted in financial gains but there was limited high quality evidence to suggest that this resulted in improved uptake or measurable health or social benefits (Adams, White et al. 2006).

Since 2010 in the UK, significant reforms made to the social security system generated confusion for those already accessing benefits, as well as those possibly entitled to them. The Welfare Reform Act 2012 legislated for Universal Credit and Personal Independence Payments (Hobson 2020). Further temporary and some more-permanent changes have been made in response to the 2020 coronavirus pandemic and continue to evolve over the course of the pandemic (Hobson 2020).

In light of the changing situation in the UK, and the increased need for financial support for vulnerable groups in the recovery from the pandemic, this paper provides a timely update of the research evidence, building upon the results of the previous 2006 review, in order to guide future policy and practice. We conducted a critical narrative systematic review to assess the health, social and financial impacts of welfare advice services co-located in health settings and to explore the facilitators and barriers to successful implementation of these services to understand how to reach those populations most in need of this service, whilst representing value for money for commissioners and society.

### OBJECTIVES

This review explores the effectiveness and experiences of welfare advice services co-located in a health setting in the UK. The objectives are to:

1. Determine the evidence of effectiveness of welfare advice services co-located in a health setting on health and social outcomes, using a meta-analysis where possible.
2. Assess the economic benefits of co-located welfare advice services from the perspective of the individual, the national health service (NHS), the commissioner and society.
3. Identify and explore the relationships between reported facilitators and barriers to implementation, to understand how and why particular barriers and/or enablers to implementation operate.

## METHODS

A critical narrative systematic review (Popay, Roberts et al. 2006) was conducted structured in accordance with recommendations from the Preferred Reporting Items for Systematic Reviews and Meta-Analyses (PRISMA) guidelines (Shamseer, Moher et al. 2015). A narrative synthesis was conducted as a method to synthesise research in the context of the systematic review. It adopts a narrative summary of the findings, alongside a statistical analysis, in order to support the process of synthesis (Rodgers, Sowden et al. 2009). This approach was chosen in anticipation of fewer empirical studies and a high volume of qualitative research and grey literature, based on the previously conducted systematic review and an initial scoping search.

### SEARCH STRATEGY

A literature search was conducted for relevant published articles from the sources listed in Table 1. Search strategies were developed, built upon the previous systematic review in this area (Adams, White et al. 2006), separately for each of the academic databases, in order to match the appropriate indexing terms, see Appendix One. The search results were limited to those written in English with a publication date between January 1^st^, 2010 and 30^th^ November 2020.

**Table 1.** Literature sources searched for studies of the health, social and financial effects of welfare services co-located in health settings

| Electronic databased searched | Websites | Other sources |
| --- | --- | --- |
| ▪ Applied Social Sciences Index and Abstracts (ASSIA)<br>▪ Humanities Index<br>▪ Cumulated Index to Nursing and Allied Health Literature (CINAHL)<br>▪ Cochrane Database of Systematic Reviews<br>▪ EMBASE<br>▪ Health Management Information Consortium<br>▪ International Bibliography of the Social Sciences (IBSS)<br>▪ Medline<br>▪ NHS Economic Evaluation Database<br>▪ PAIS Index<br>▪ Psycinfo<br>▪ Science Citation Index<br>▪ Social Policy and Practice and Social Care Online<br>▪ Social Science Citation Index<br>▪ Social Services Abstracts<br>▪ Sociological Abstracts<br>▪ Taylor & Francis<br>▪ WorldCat<br>▪ Zetoc | ▪ Age Concern<br>www.ageconcern.org.uk<br>▪ Child Poverty Action Group<br>www.cpag.org.uk<br>▪ Department of Health (UK)<br>www.dh.gov.uk<br>▪ General Accounting Office (US) www.gao.gov<br>▪ Home Office (UK)<br>www.homeoffice.gov.uk<br>▪ Joseph Rowntree Foundation<br>www.jrf.org.uk<br>▪ MDRC www.mdrc.org<br>▪ National Audit Office (UK)<br>www.nao.org.uk<br>▪ Office of Policy (US)<br>www.ssa.gov/policy<br>▪ Office of the Deputy Prime Minister www.odpm.gov.uk<br>▪ Rightsnet<br>www.rightsnet.org.uk<br>▪ Urban Institute<br>www.urban.org | ▪ Hand searching of key journals<br>▪ Google<br>▪ Google Scholar<br>▪ Reference list of included articles<br>▪ Author searches<br>▪ Conference publications<br>▪ Published policies<br>▪ Other relevant grey literature |

### INCLUSION AND EXCLUSION CRITERIA

This systematic review includes studies which qualitatively or quantitatively examined the impact of welfare advice services delivered whilst physically co-located in a health setting in the United Kingdom, on any outcome (including health, social, financial outcomes), published from January 2010 to November 2020.

Studies published outside the UK were excluded, given the significant variation in nature, provision and funding of both welfare services and healthcare settings. Studies published prior to 2010 were also excluded, owing to the significant reforms made to the social security system in the UK. Moreover, studies examining the provision of specialist services (e.g. housing advice for homeless people) were excluded from the study, as these services are not delivered as general welfare advice services by welfare advisors.

For the purposes of this review, healthcare settings are those defined as health care related buildings, where the primary purpose is to promote, restore or maintain health (World Health Organisation 2009). Welfare advice services are defined as the delivery of expert advice concerning general welfare rights and entitlement to and claims for welfare benefits.

### DATA EXTRACTION AND RECORD MANAGEMENT

Following completion of the literature search, the results were exported to Covidence (Veritas Health Innovation). Screening was performed through a process of marking records for inclusion based on the relevance of the title, followed by the abstract and full text. The accuracy of the selection was checked by a second reviewer who repeated the abstract and full text selection process independently with a random sample of 10% of excluded studies.

Data were extracted using a structured, pre-piloted, proforma using Covidence software. Headings adapted from Popay et al. were used to structure the data extraction: setting, participants, aim, sampling and recruitment, method, analysis and results (Popay, Roberts et al. 2006). The reference management software, EndNote, was used to store and manage the retrieved references.

### QUALITY ASSESSMENT

The quality and risk of bias of each study was assessed using tools from the Center for Evidence-Based Management (CEBMa) according to study design (Center for Evidence-Based Management 2014). The CEBMa does not include a tool for studies adopting a mixed methods design. For these studies, the Mixed Methods Appraisal Tool (MMAT) was used (Hong, Fàbregues et al. 2018). Studies were assessed based on the clarity of the research question, eligibility criteria, study population and sample size, outcomes measured, and type of statistical analysis employed.

After evaluation, studies were classified into three appraisal categories (high, medium and low) based on their internal validity indicated by the quality appraisal score.

Alongside a quality and risk of bias assessment, all studies were appraised using tools to evaluate the relevance and ‘richness’ of their findings. ‘Richness’ has been described as ‘the extent to which study findings provide in-depth explanatory insights that are transferable to other settings’ (Popay, Roberts et al. 2006). The criteria for assessment of ‘richness’ taken from an approach by Higginbottom et al. (2020) are described in Table 2 (Higginbottom, Morgan et al. 2013, Higginbottom, Evans et al. 2020).

**Table 2.**
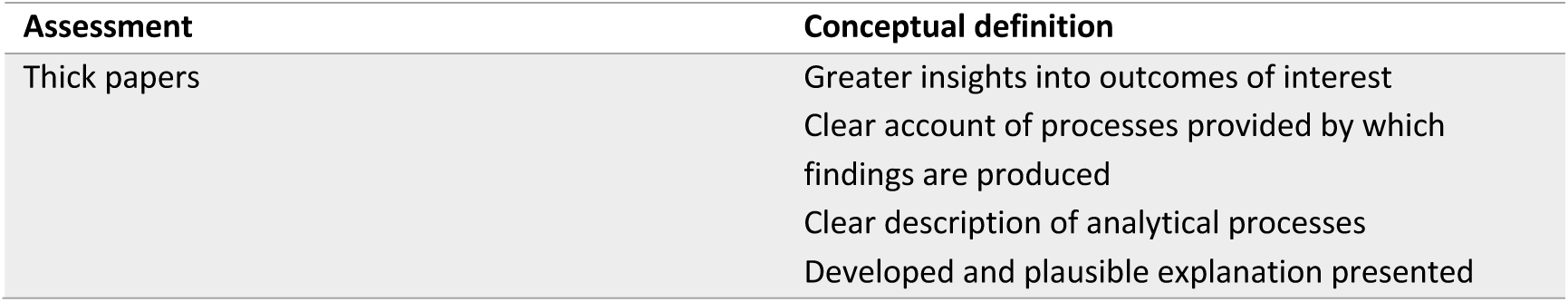
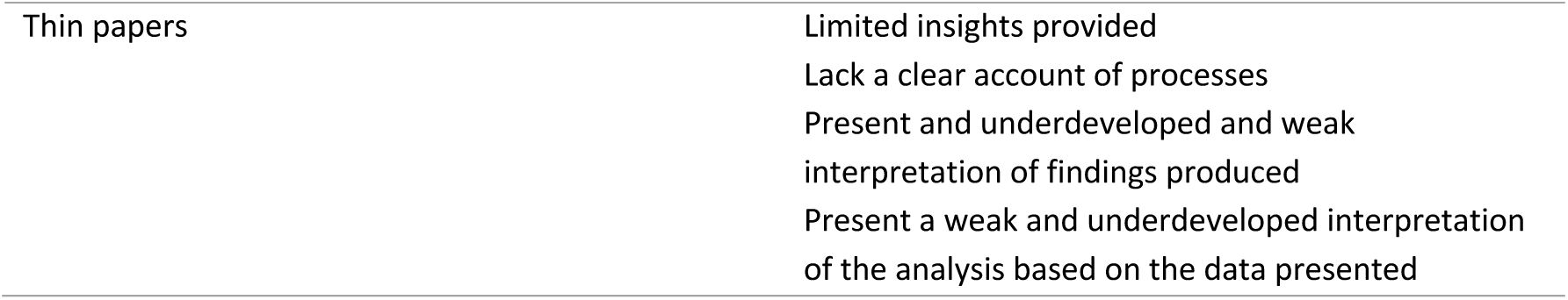
Criteria for assessment of ‘richness’ (Higginbottom, Morgan et al. 2013, Higginbottom, Evans et al. 2020)

| Assessment | Conceptual definition |
| --- | --- |
| Thick papers | Greater insights into outcomes of interest<br>Clear account of processes provided by which findings are produced<br>Clear description of analytical processes<br>Developed and plausible explanation presented |
| Thin papers | Limited insights provided<br>Lack a clear account of processes<br>Present and underdeveloped and weak interpretation of findings produced<br>Present a weak and underdeveloped interpretation of the analysis based on the data presented |

### DATA SYNTHESIS

Data extracted from the included studies were analysed using a critical narrative synthesis, adopting an evidence-led framework described by Rodgers et al. (Rodgers, Sowden et al. 2009). This approach consists of four elements employed in an iterative manner to analyse the included studies: developing a theory of how the intervention works, why and for whom; developing a preliminary synthesis of findings of included studies; exploring relationships in the data; and assessing the robustness of the synthesis.

An overarching theory of change was developed a priori and will be presented to explain how the intervention works, why and for whom. This was used as an analytical framework against which to assess the evidence and will be refined in light of the emerging findings from the narrative systematic review. This was achieved using a theory of change model which describes ‘the chain of causal assumption that link programme resources, activities, intermediate outcomes and ultimate goals’ (Brannen 2005, Mayne 2015).

A textual description of all included and excluded studies was created alongside the quality assessment to generate summary measures that were used to form a cross-study analysis. An example one page systematic textual narrative summary can be found in Appendix X (Popay, Roberts et al. 2006, Higginbottom, Evans et al. 2020).

Where the results from studies could be sensibly pooled, this was conducted using appropriate meta-analytic techniques. Where significant heterogeneity of the study methodology, or lack of formal statistical analysis, was identified, quantitative data are presented descriptively. The average values across the studies are reported, alongside the median and range where appropriate, to give an indication of spread and variability of data.

Qualitative data were translated through a thematic analysis, chosen for its systematic and replicable approach to analysis based on explicit rules of coding (Stemler 2000). The data were interrogated to explore relationships within and across the included studies. Factors were identified that might explain differences in direction and size of effect across the included studies or in the type of facilitators and/or barriers to successful implementation of co-located welfare rights advice interventions.

Heterogeneity between all studies was explored in consideration of study design, outcomes and study population. Given the complex nature of welfare rights advice interventions, it was difficult to anticipate the main sources of heterogeneity a priori. Where the main potential sources of variation could be identified, heterogeneity between effects were explored by means of subgroup analysis, based on the theory of change model about how the intervention works and for which groups.

Sub-group analyses and analyses of moderator variables were used to explore the effects of the interventions within and between studies, using variability in population characteristics and referral mechanism as modifiers, along with any other modifiers highlighted during analysis. Where appropriate conceptual models and concept mapping were used to explore and highlight relationships between data.

### THEORY OF CHANGE

Our theory of change proposes that the implementation of a welfare advice service in a health setting results in cost savings to the NHS and social sector and ultimately reduces health inequalities, see Figure 1. This was used as an analytical framework against which to assess the evidence and will be refined in light of the emerging findings from the narrative systematic review.

**Figure 1.**
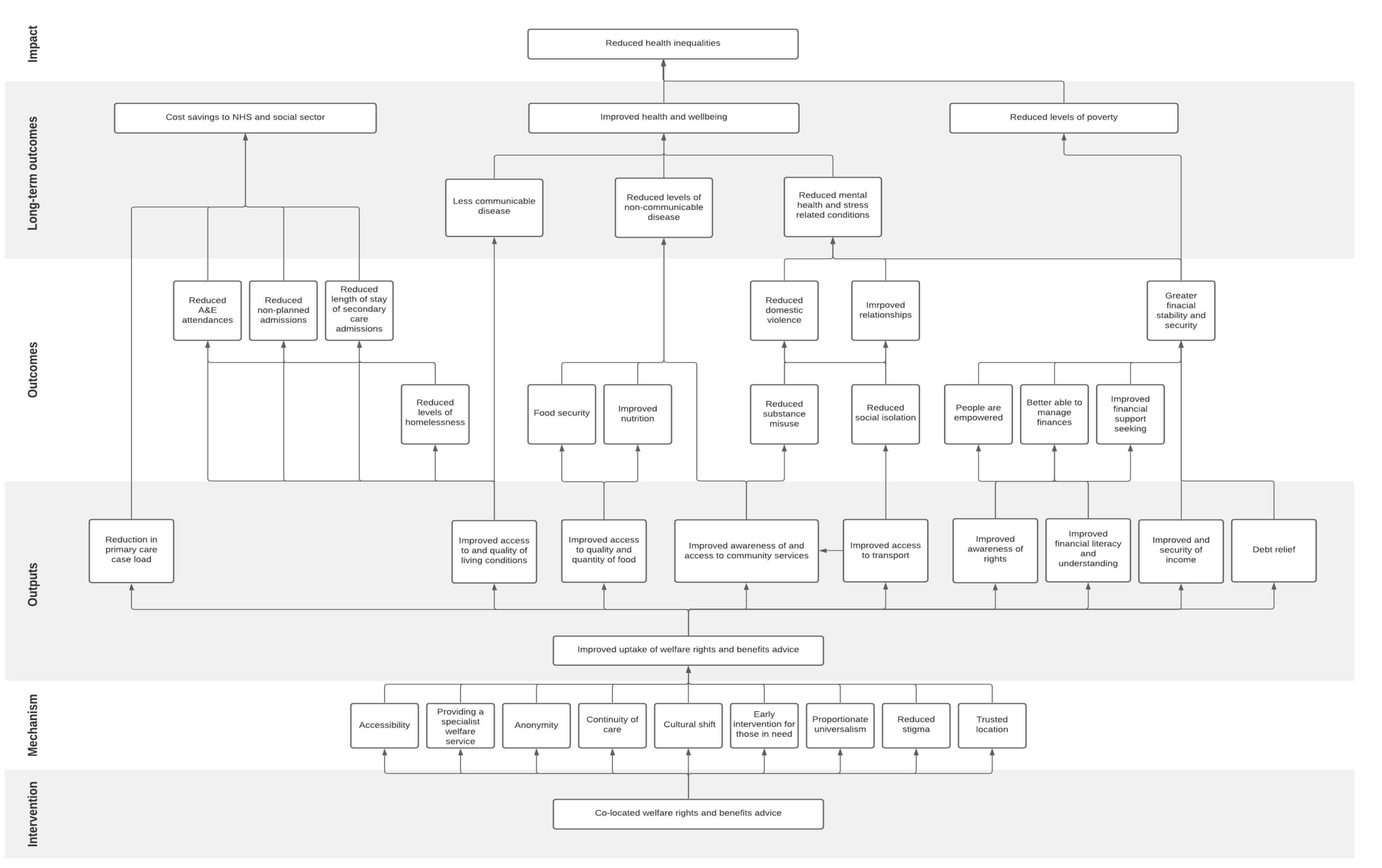
Theory of change model to propose how the implementation of a welfare advice service in a health setting can results in cost savings to the NHS and social sector and ultimately reduce health inequalities.

There are several mechanisms through which welfare advice services co-located in a health setting might operate to improve uptake of advice, compared to welfare advice services offered in a conventional setting, owing to the nature of its co-location. Being nested within a health setting, the services are considered more accessible and provide a greater degree of anonymity to individuals accessing them. Due to the connection between welfare advice services and health professionals, the services are perceived to be more trustworthy, less stigmatising and better able to identify and provide early intervention to those most in need of help. The services are thought to offer a more enhanced, specialist service, tailored to the needs of those specifically with long-term health and mental health conditions, with better follow up and continuity of care, compared to conventional services. Overall, welfare services co-located in a health setting adopt a proportionate universalism approach, distributing resources to favour the disadvantaged, by increasing resources to meet the needs of some of society’s most vulnerable people, enabling it to have a greater impact on health inequalities (Mayne 2015).

Access to these services and take up of the welfare advice provided, improve financial security and stability for individuals through increased household income and support with debt relief. Improved financial literacy and an awareness of their welfare rights, help individuals feel more empowered and better able to manage their finances and improves their financial support seeking when they are in need of financial assistance in the future, instead of relying on overdrafts, credit cards and loans. This breaks the cycle of spiralling financial insecurity and ultimately reduces levels of poverty. These impacts on financial security improve physical health and wellbeing, through reduced levels of mental health and stress-related conditions.

Accessing co-located welfare services could also improve health and wellbeing through measures to address other social determinants of health more directly. The services provide advice and support to improve housing conditions, access to nutritional food and transport, reducing the risk of communicable disease transmission and improving physical health, as well as mental health and wellbeing. Services also raise awareness of and promote access to community services, improving and encouraging appropriate use of health services to improve health and wellbeing generally. This also reduces levels of substance misuse directly, improving personal relationships and reducing levels of domestic abuse, all improving health and wellbeing.

Finally, improved access to welfare services may also provide benefits to the NHS. Improved uptake of welfare advice services lead to a reduction in primary care appointments and improved use of secondary health services, particularly mental health services, resulting in significant cost savings for the NHS and freeing up the resources needed to address those most in need.

## Results

The search identified 7998 potentially eligible records through bibliographic database searches and an additional 15 from reference and citation searching. Upon removal of duplicates and exclusion after title and abstract review, 138 articles were left for full text review. A total of 14 studies were included in the final review, see Figure 2. A description of each included study is outlined in Table 3. Superscript references in the text will be used to refer to the relevant included studies, numbered according to their place in Table 3.

**Table 3.**
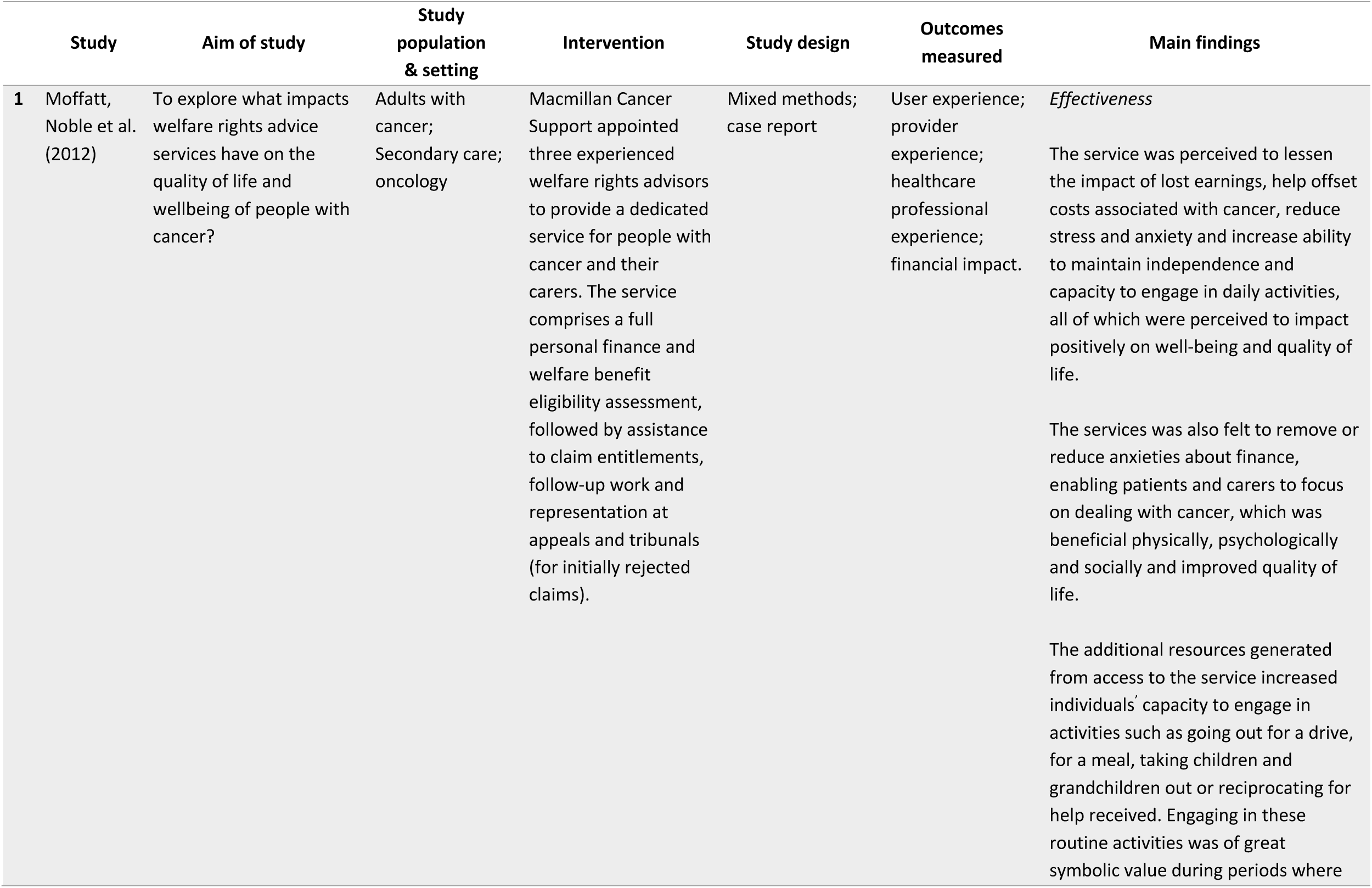
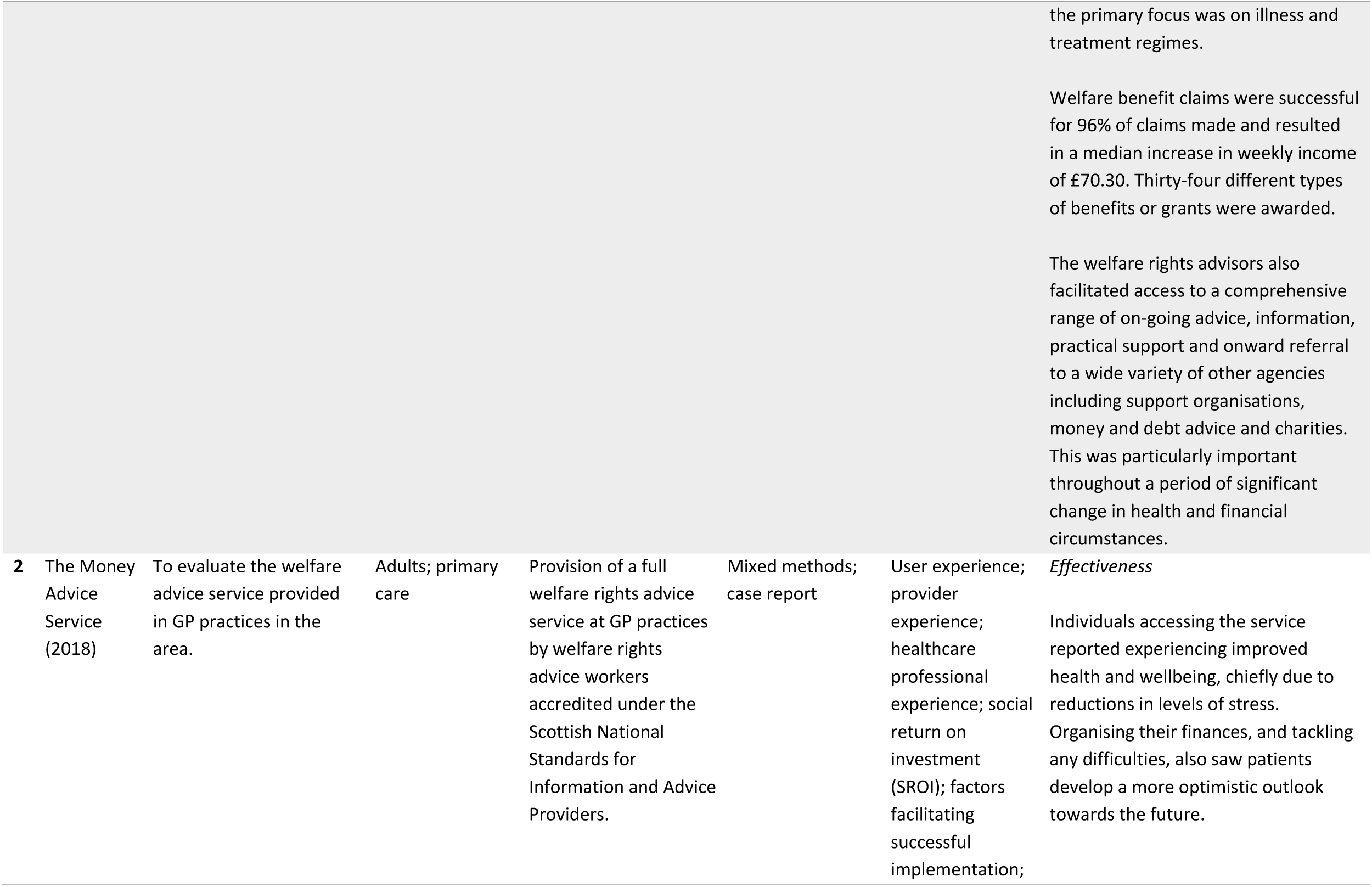
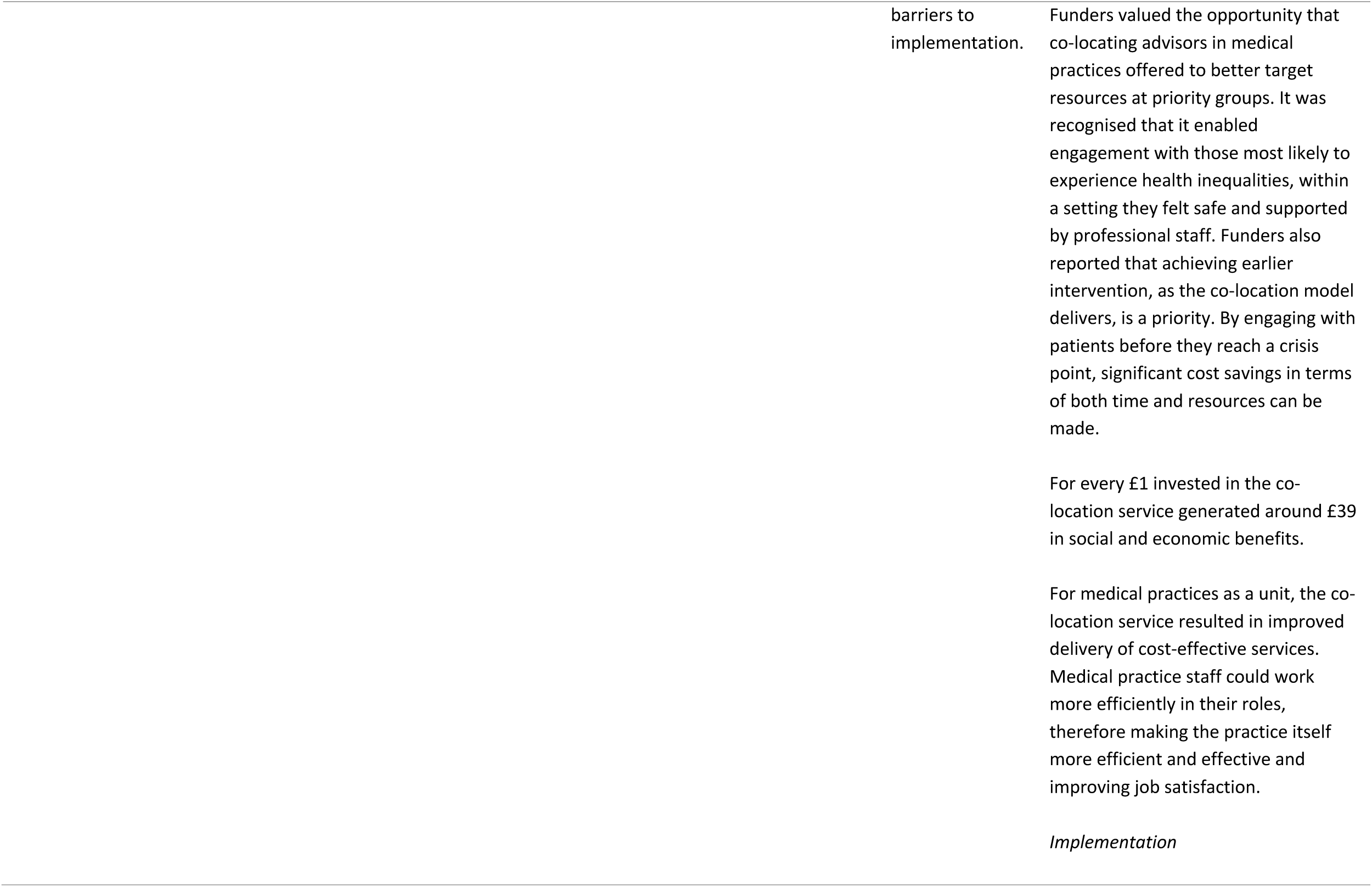
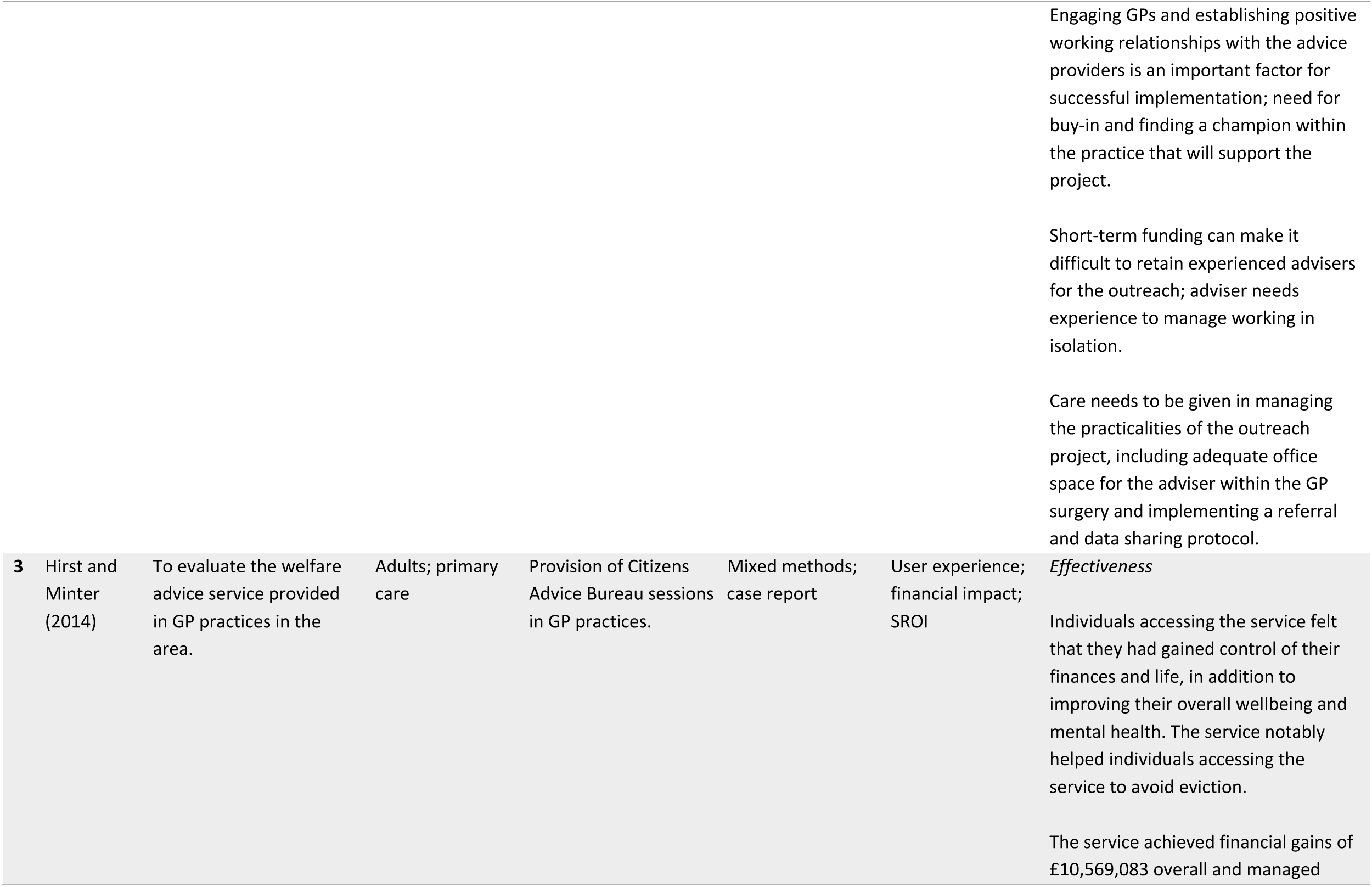
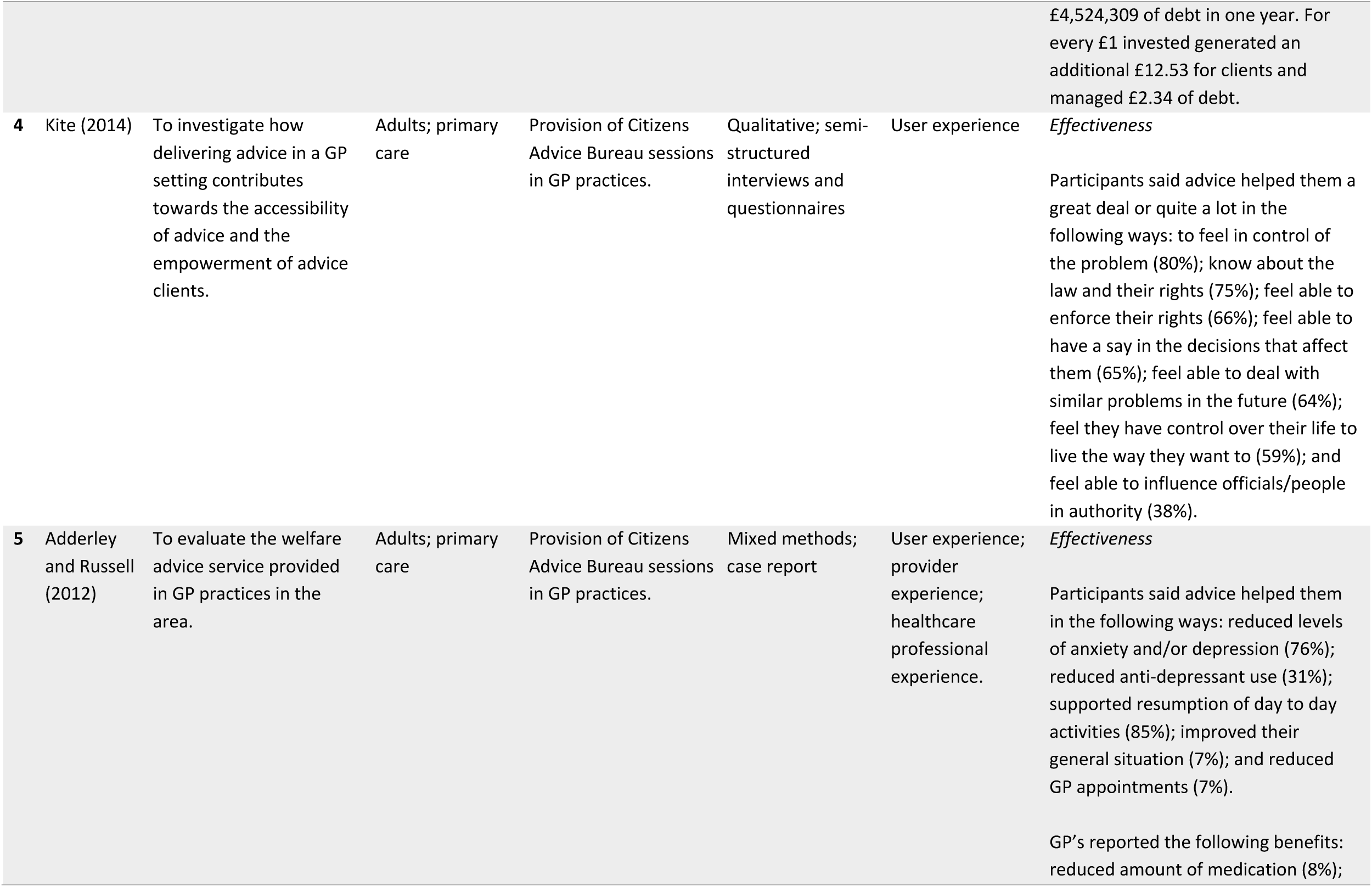
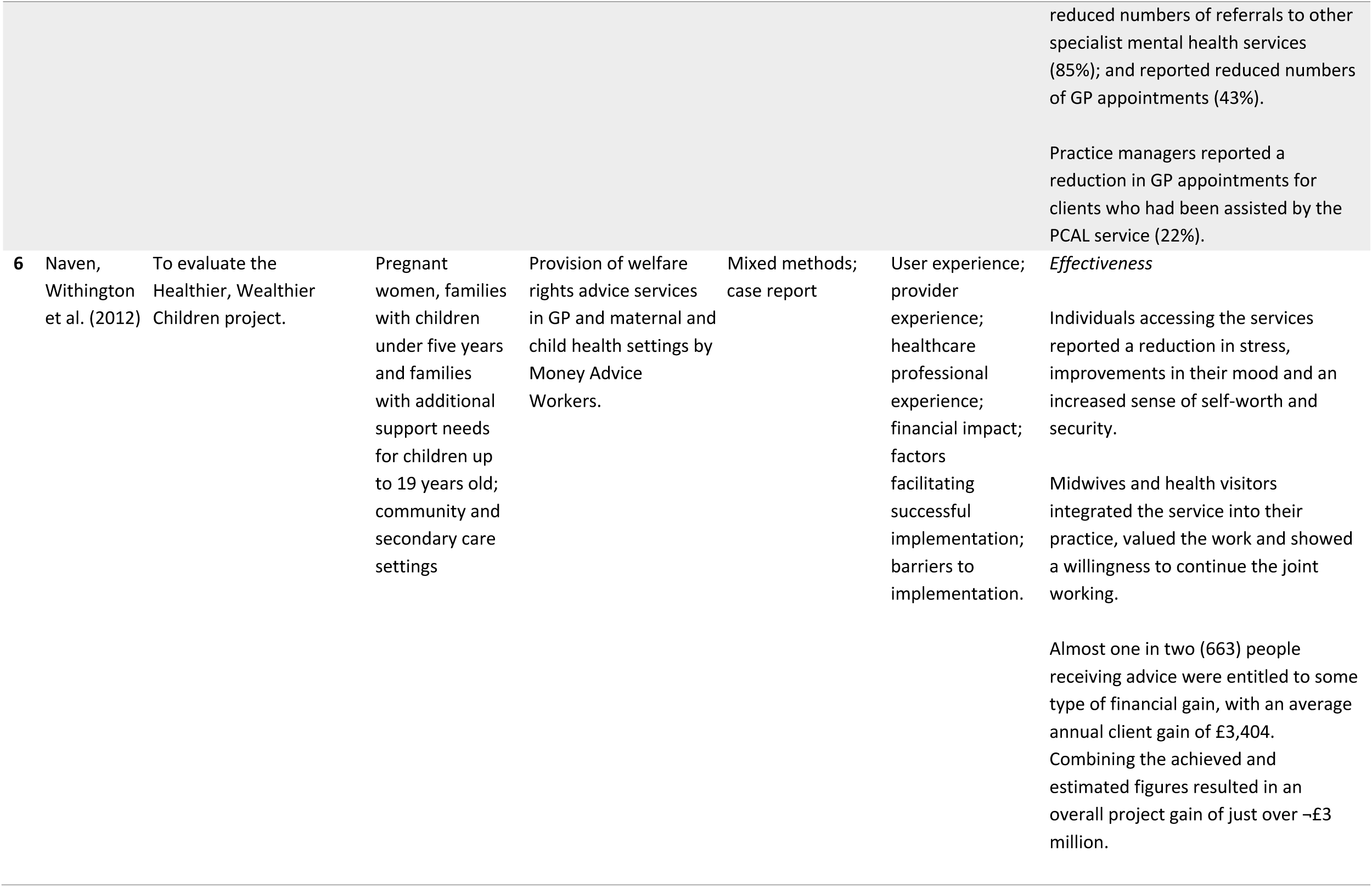
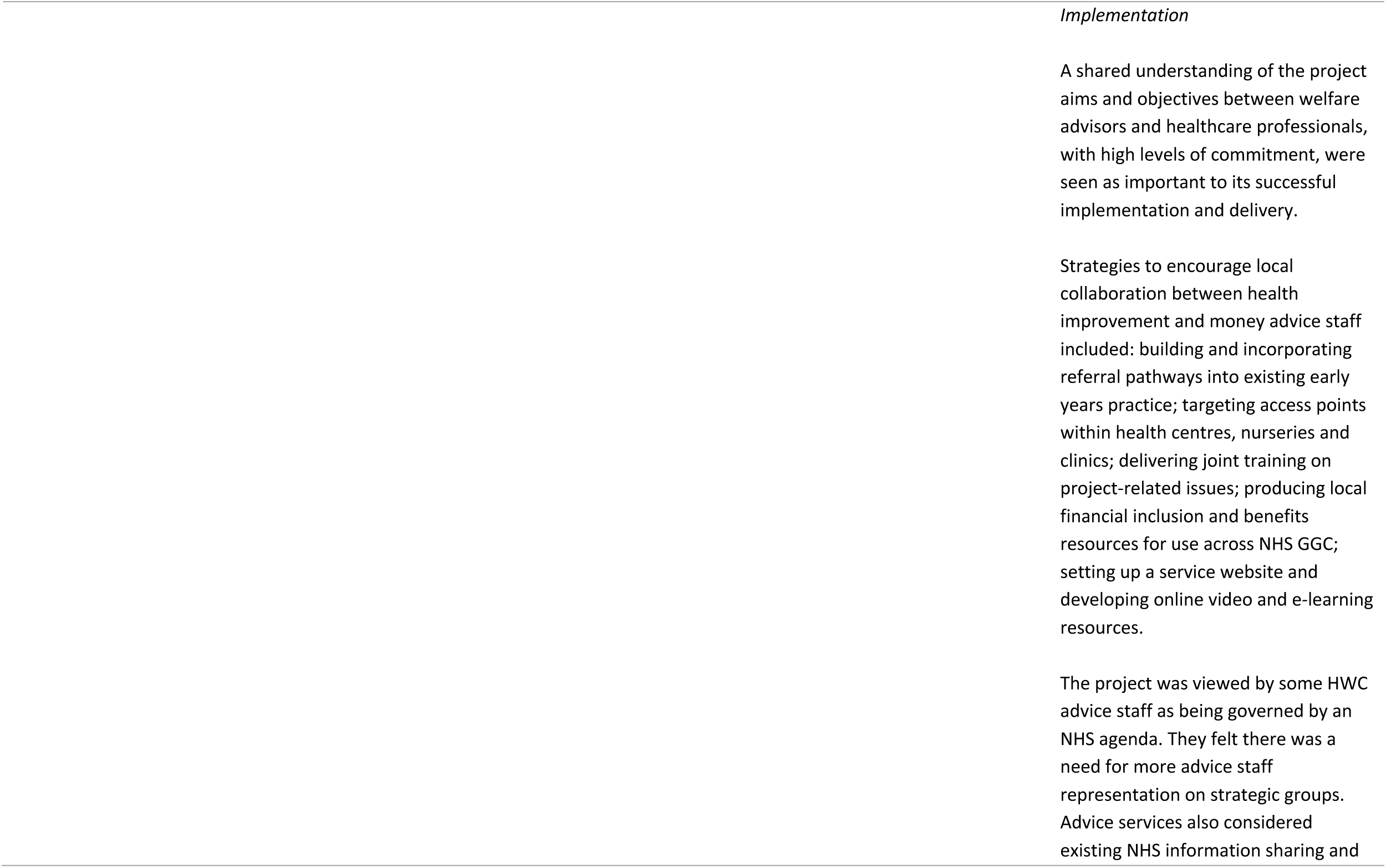
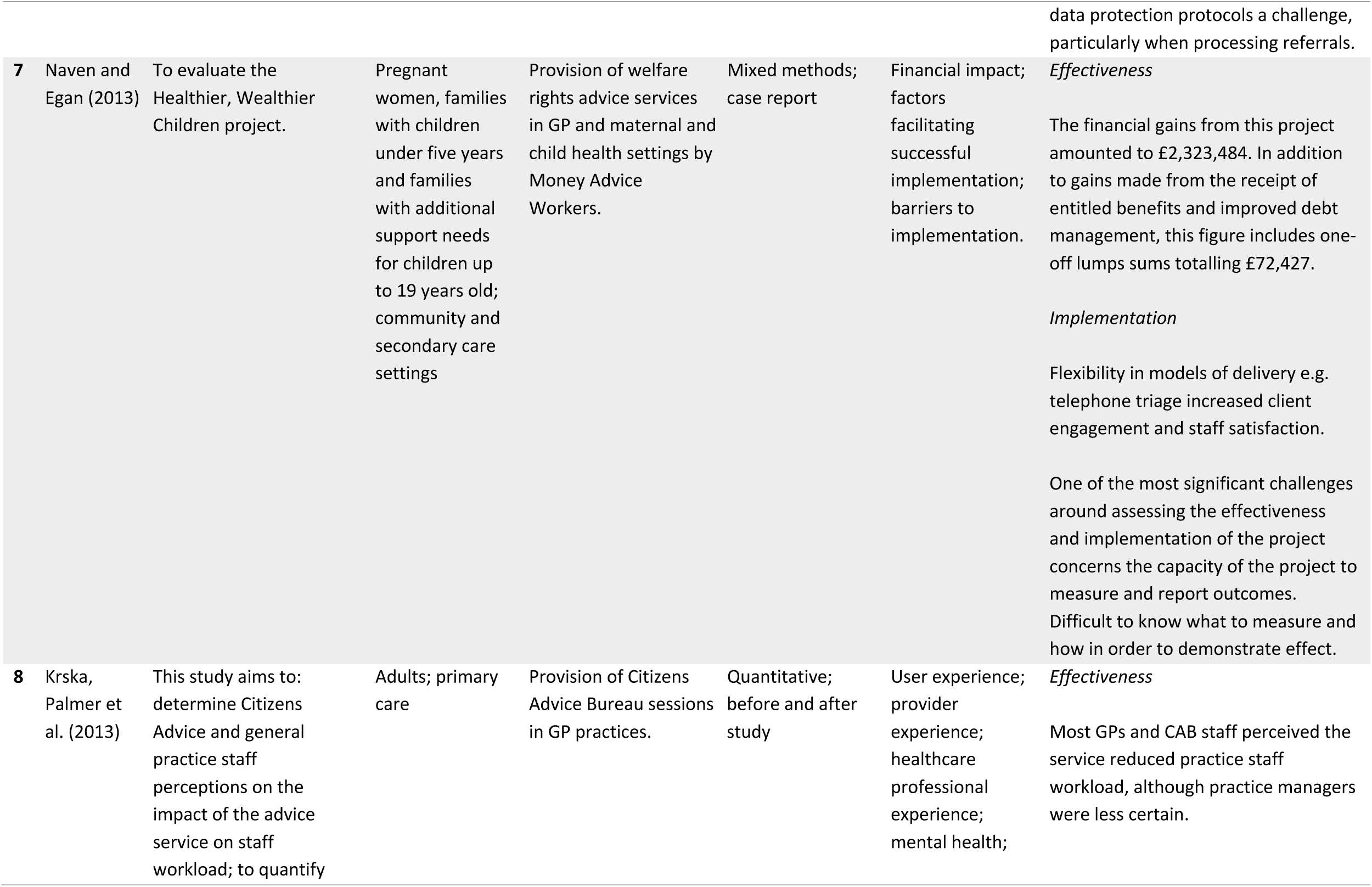
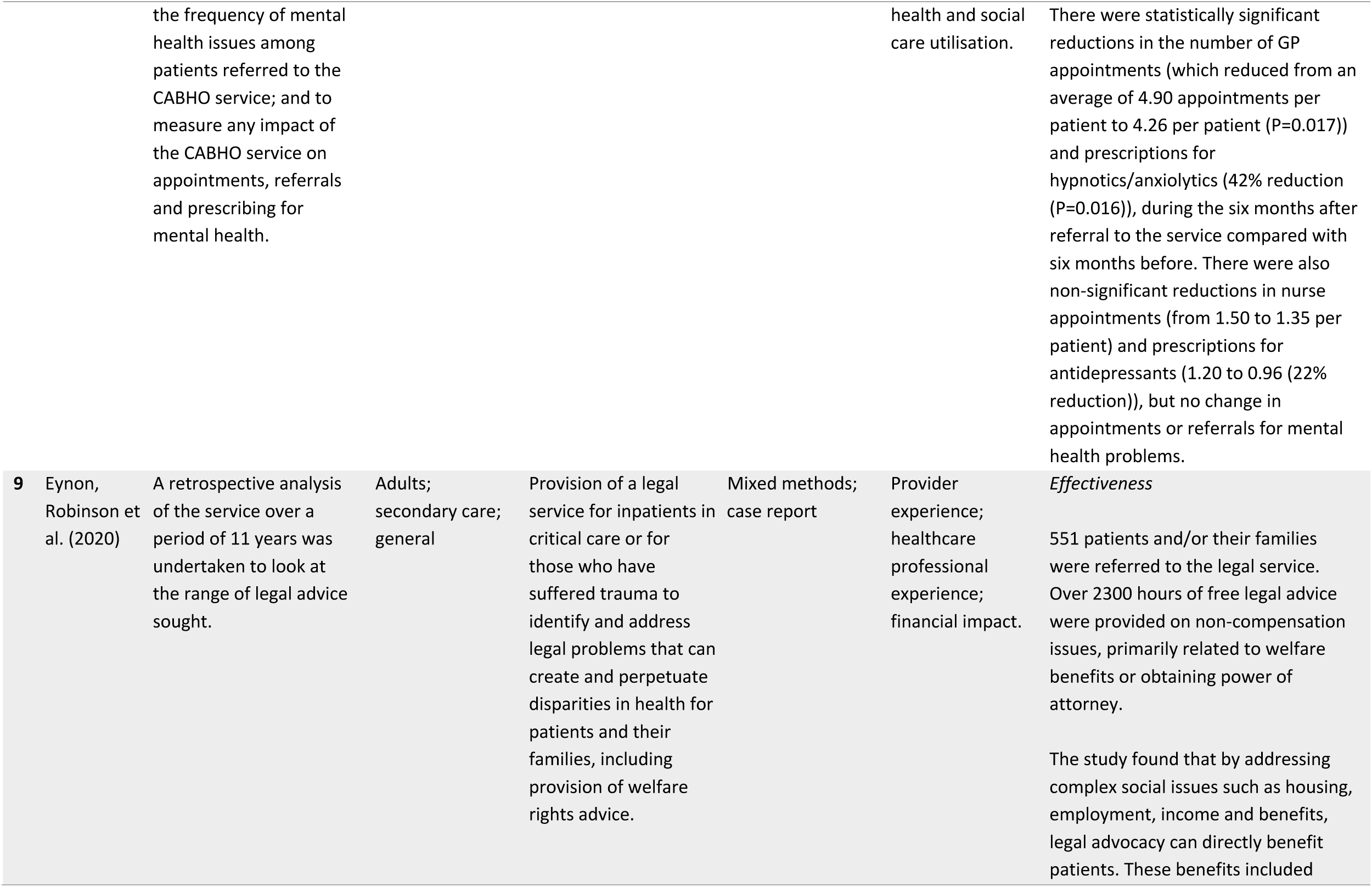
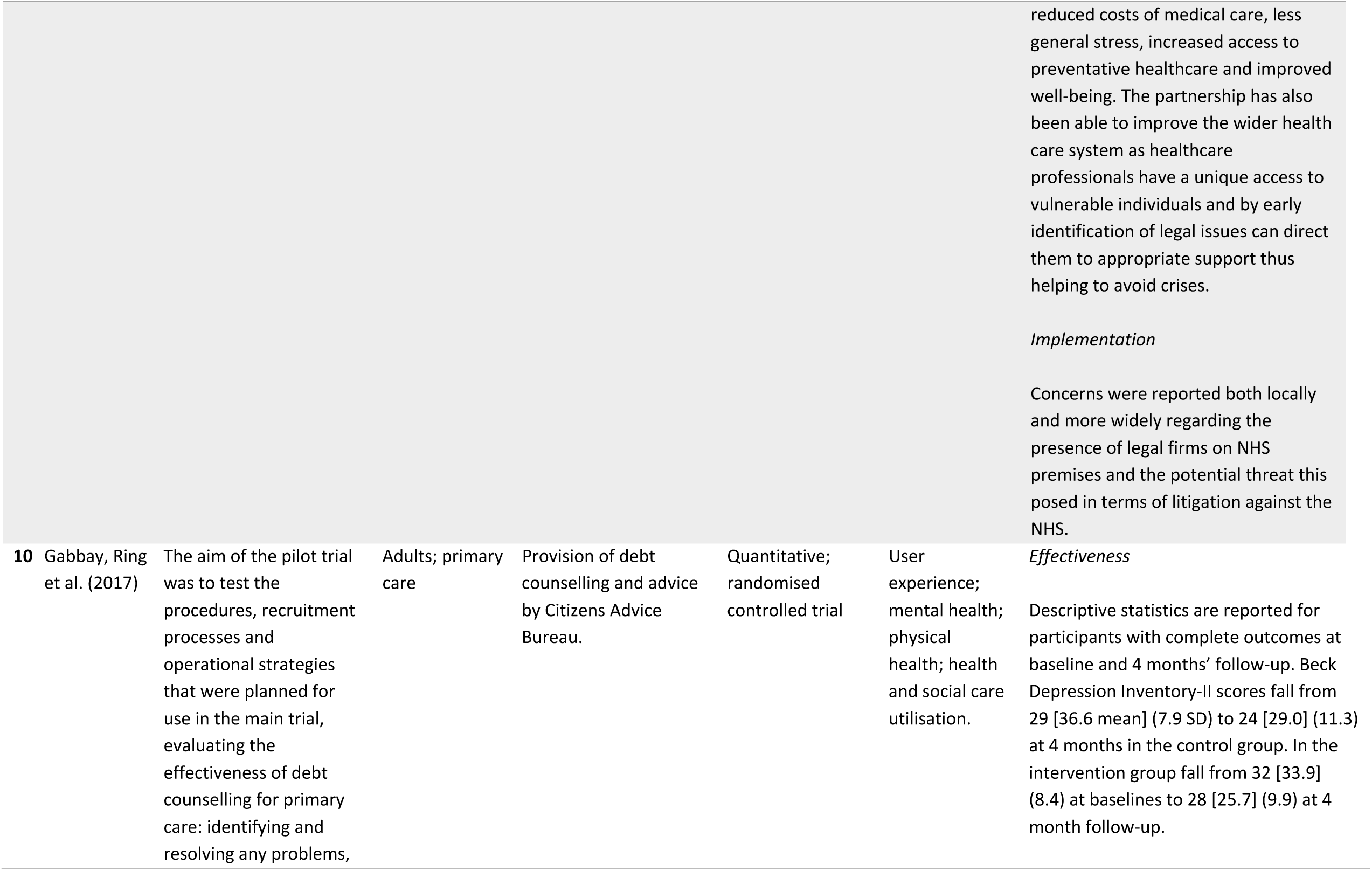
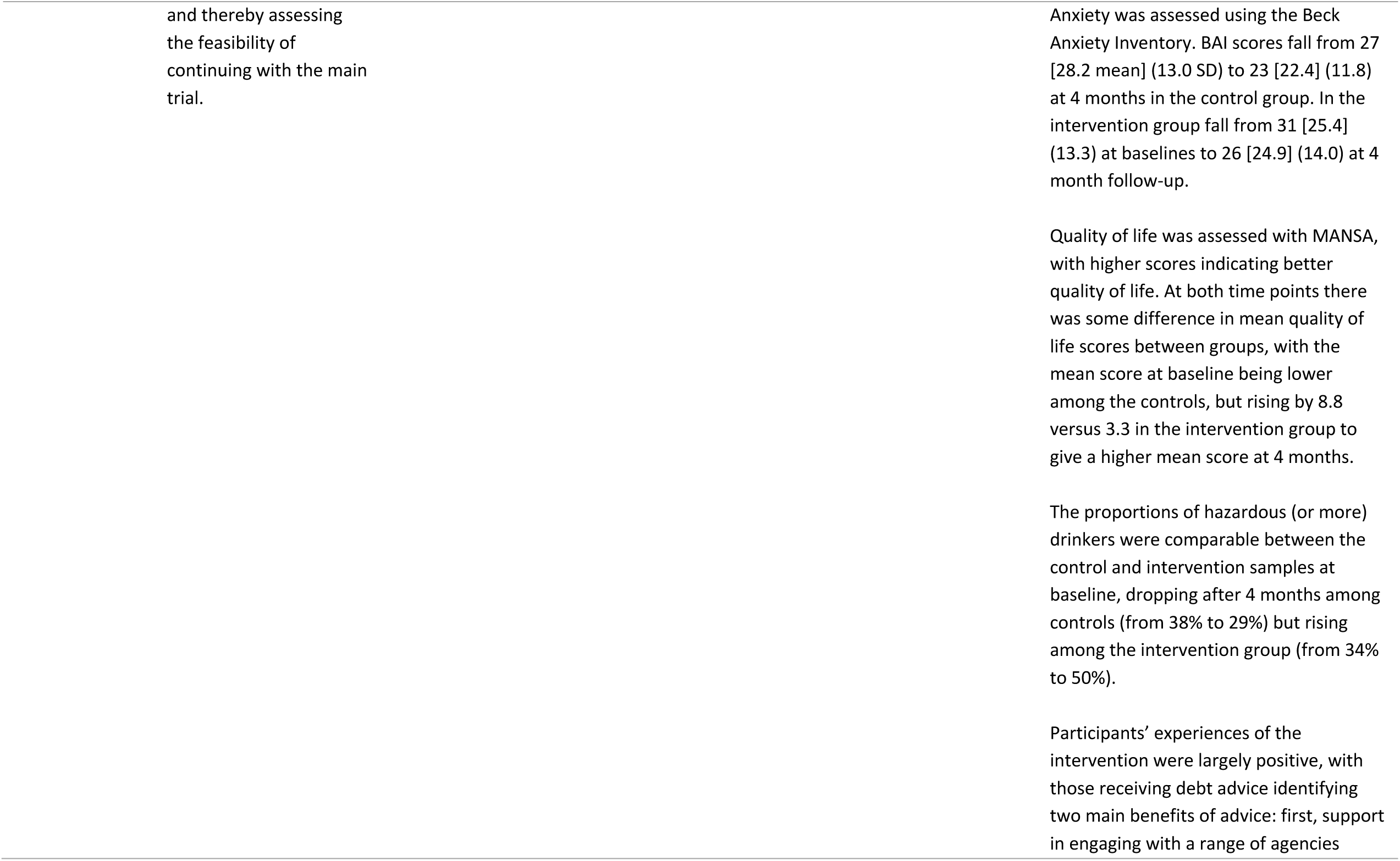
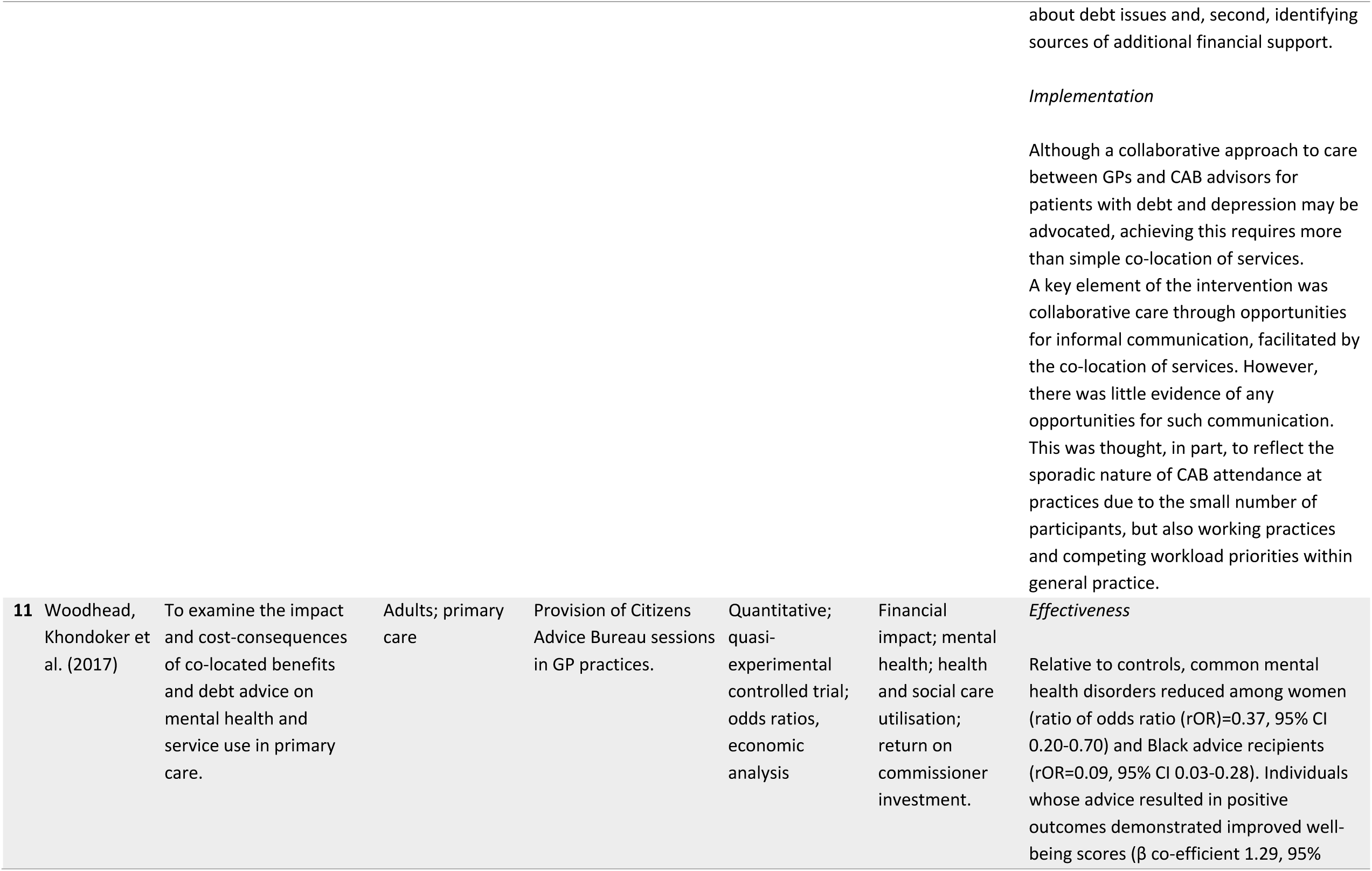
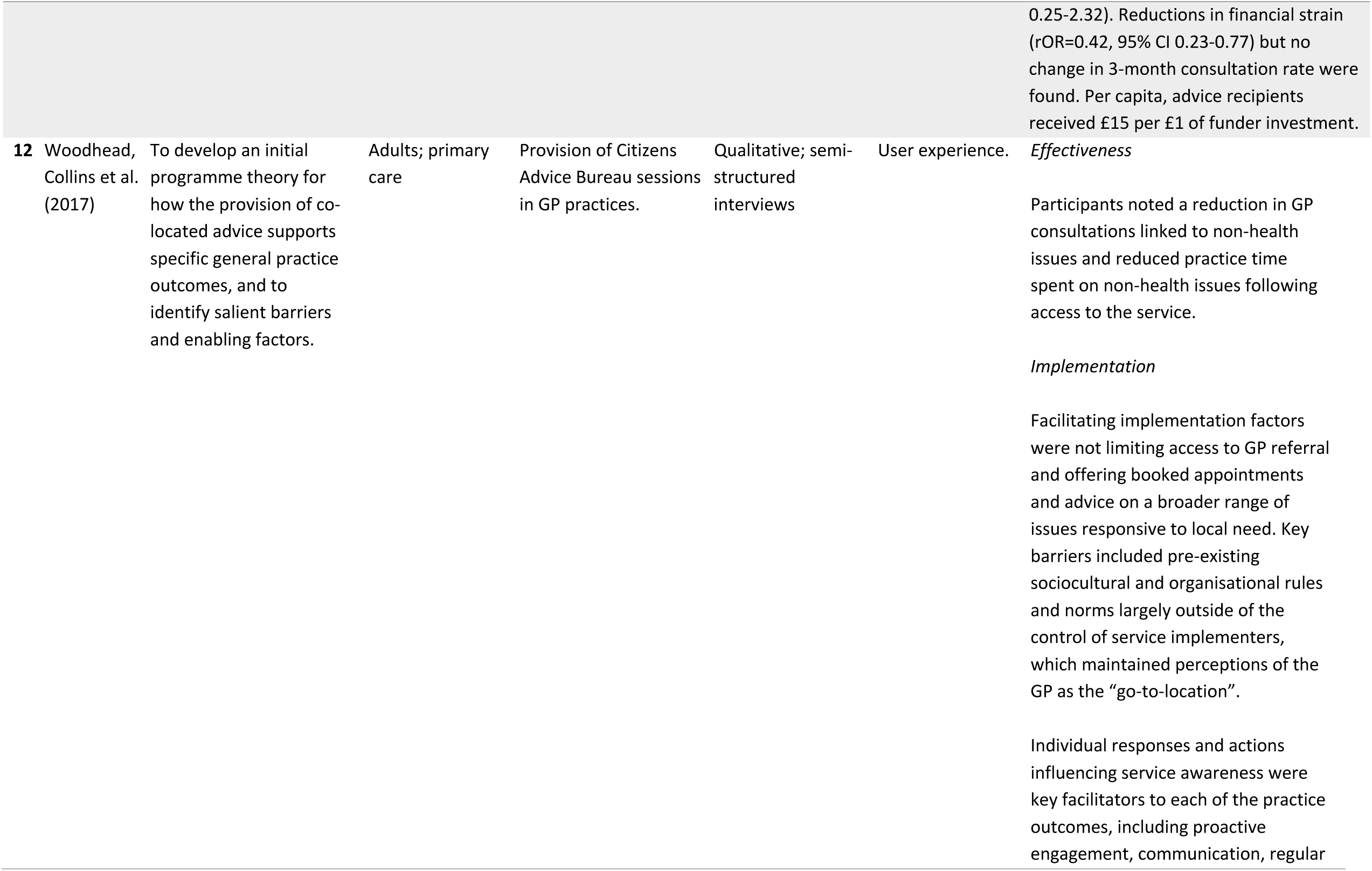
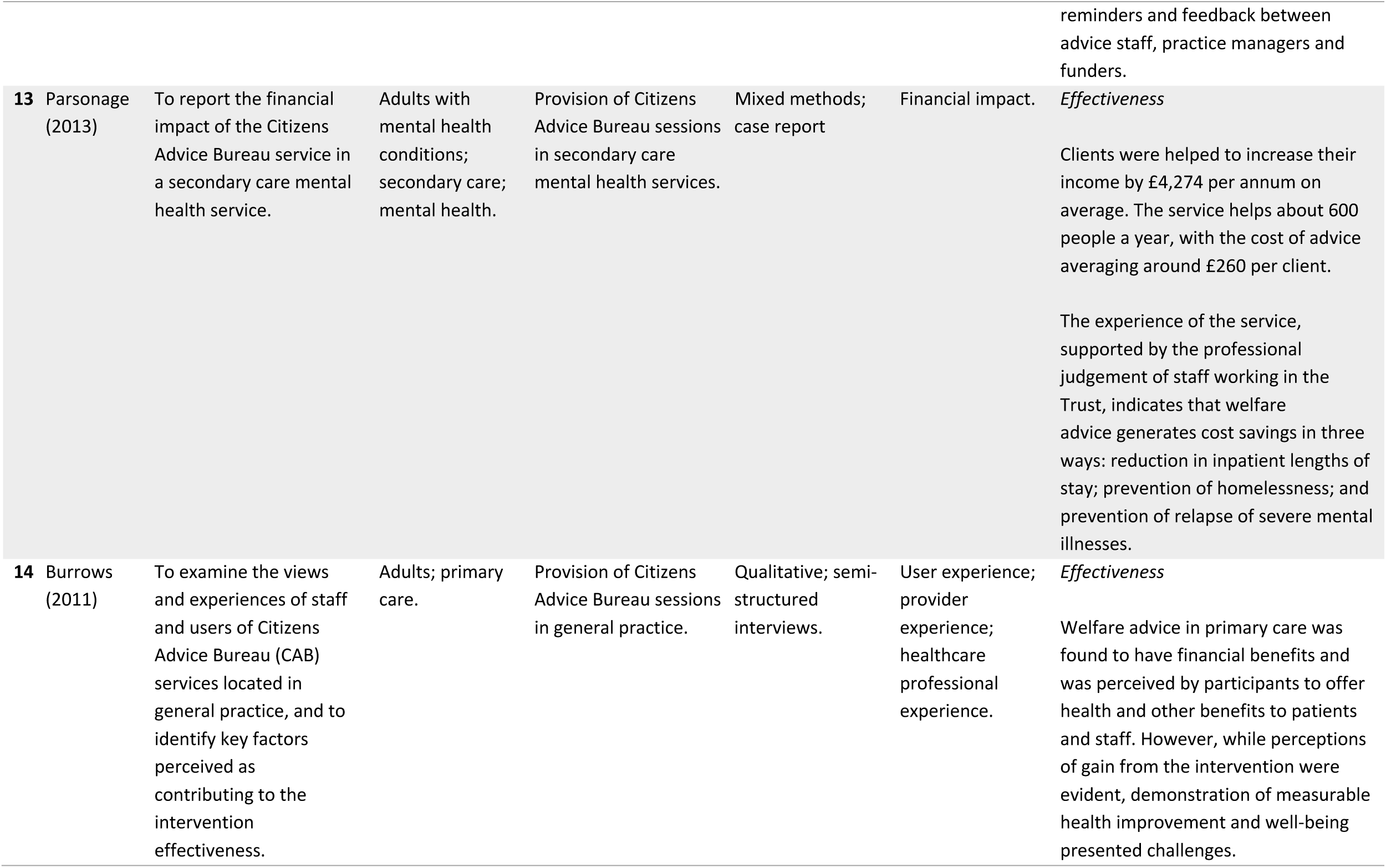
Characteristics and narrative description of included studies in the narrative systematic review.

**Figure 2.**
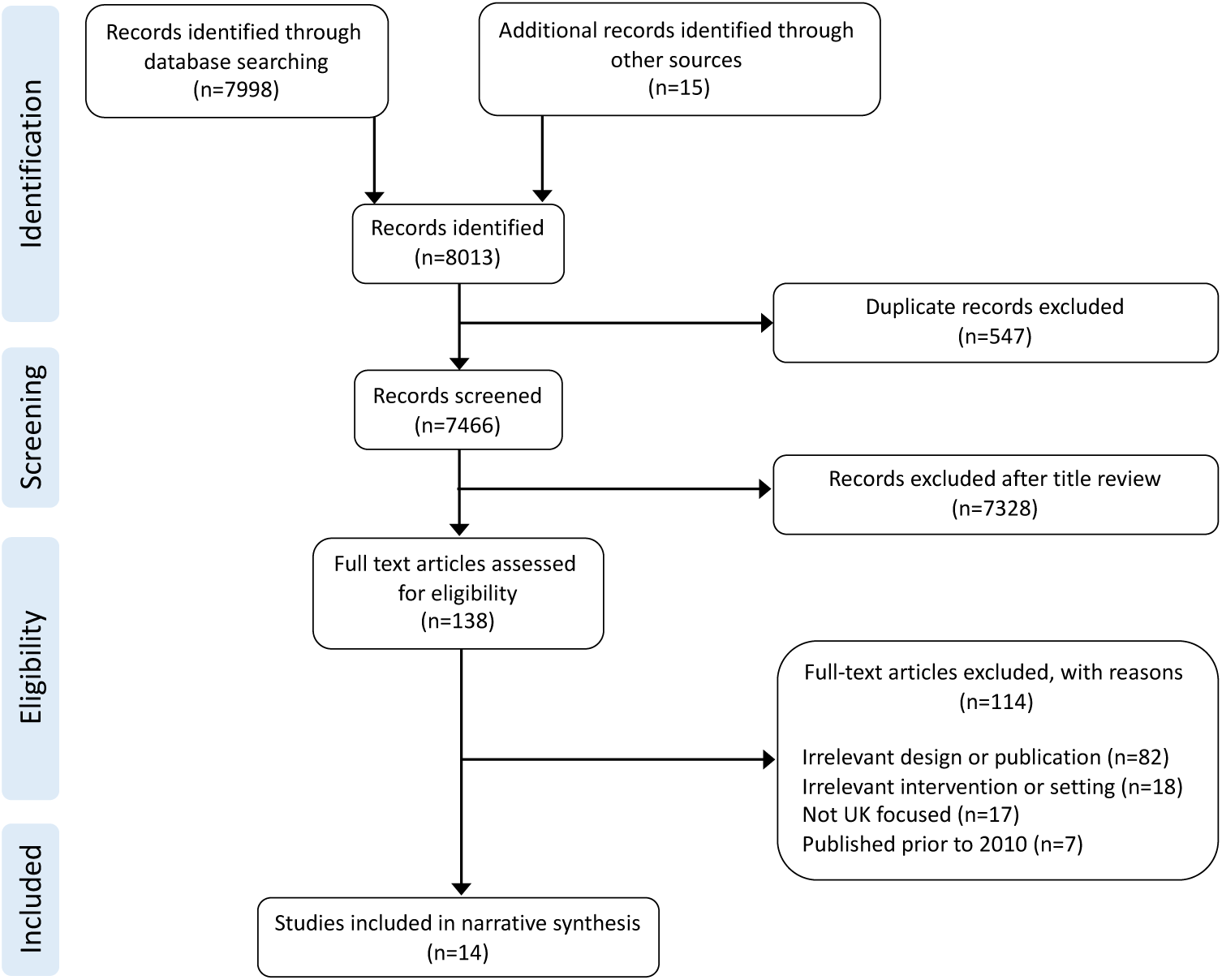
The PRISMA flow chart of the final selection process

### STUDY CHARACTERISTICS

Of the 14 studies included in this review, half were published in peer-reviewed journals,^1,8-12,14^ six studies were published as reports,^2-3,5-7,13^ and one was published as a thesis abstract.^4^ The included studies were published between 2010 and 2020, nine prior to 2015.^1,3-8,13-14^ They employed a range of designs: one non-randomised controlled trial,^11^ one pilot randomised controlled trial which was terminated as a result of low recruitment,^10^ one before-and-after-study,^8^ three qualitative studies, ^4,12,14^ and eight descriptive case studies.^1-3,5,6-7,9,13^ The evidence from this review has been mapped onto the theory of change model (Figure 3), demonstrating the spread of evidence across the model; highlighting areas with a greater evidence base and areas where evidence is limited or lacking.

**Figure 3:**
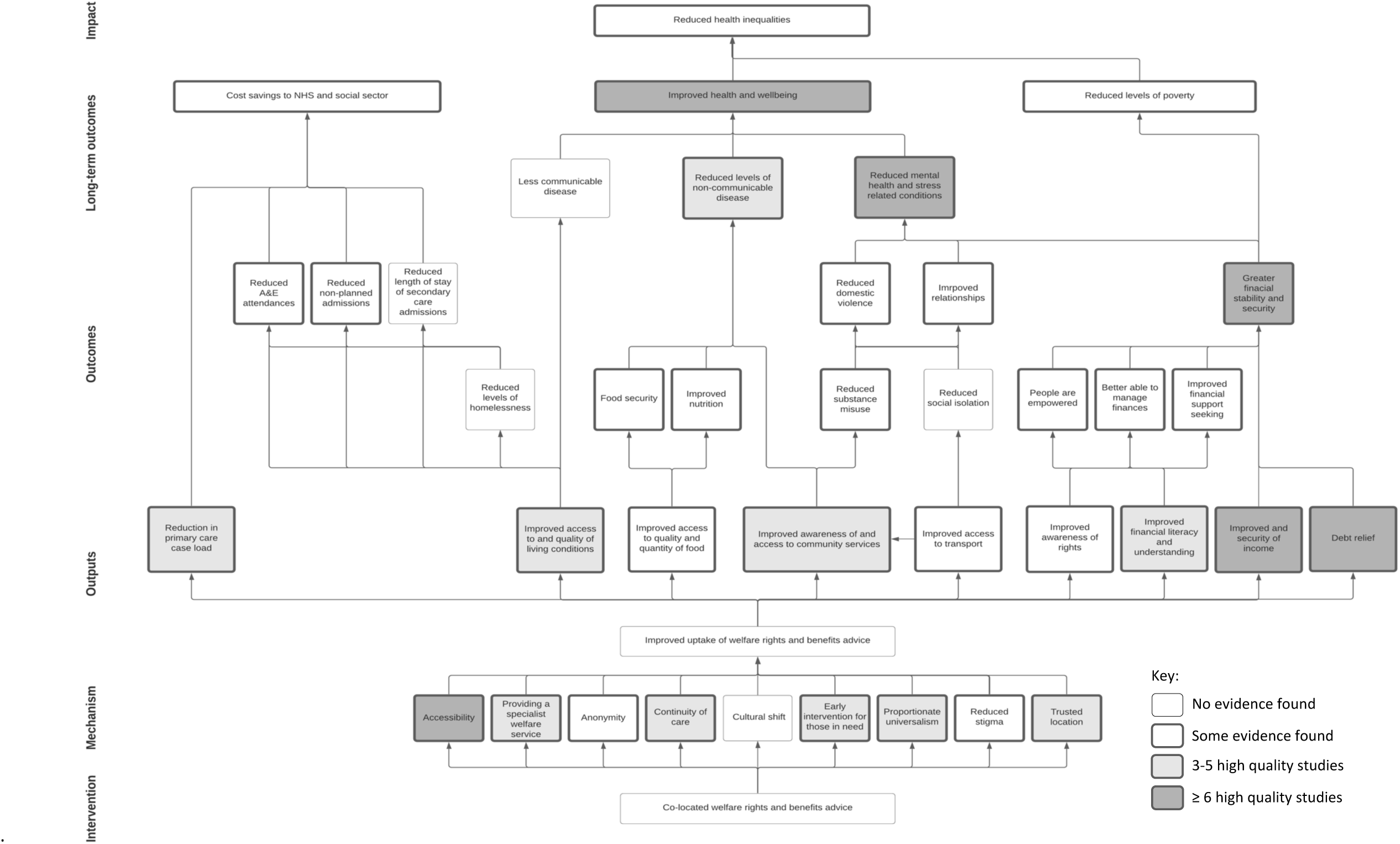
Map of the narrative systematic review evidence against the theory of change model

The welfare advice services evaluated in the reviewed studies all provided general welfare rights advice for adults aged 18 years and over, 11 were for the general population and three provided services specifically for: adults with cancer;^1^ mental health problems;^13^ or mothers and their families^6-7^. Nine of the evaluated services were co-located in general practice,^2-5,8,10-12,14^ while three were co-located in secondary care in mental health,^13^ oncology^1^ and intensive care^9^ settings. Two linked studies evaluated services co-located across maternal and child health community and secondary care settings.^6-7^ Welfare advice services co-located in a primary care setting usually provided advice and support to the general practice patient list, although some offered this more generally to the local population, not limited to those registered with the practice. Access to welfare services was largely appointment based and accessed through referral by a general practitioner. However, some patients could self-refer. Two providers offered a drop-in service.

The co-located welfare advice services were largely provided by the Citizen’s Advice Bureau (n=9) including all of those co-located in general practice in England.^3-5,8,10-14^ For services based in Scotland (n=3), the services were provided by Money Advice Workers^6-7^ or welfare advisors accredited under the Scottish National Standards for Information and Advice Providers.^2^ The co-located oncology welfare advice was provided by Macmillan Cancer Support^1^ and the welfare advice service co-located in intensive care was provided by trained legal advisors.^9^

The majority of reviewed studies reported the effects of the intervention on health^1-3,5-6,8-12,14^ (n=11) and social outcomes^1,3,6,9,11-14^ (n=8) for the participants. Three papers^8,10,11^ utilised quantitative methods and eight papers^1-3,5-6,12-14^ used forms of qualitative methods to explore physical and mental health outcomes. Social outcomes included improved access to housing, employment and education opportunities and improved relationships. Seven papers^1,3,6,9,12-14^ utilised qualitative methods and one paper^11^ used forms of quantitative methods to explore physical and mental health outcomes. Three studies reported predominately on the impact of the intervention on mental health outcomes.^8,10,13^ Six of the studies evaluated the impact of the intervention on health services, in particular its effect on prescribing, service use and staff workload.^2,5,8,11-14^

Seven studies incorporated an economic evaluation, six reporting from the perspective of the welfare advice recipient,^1-3,6-7,11,13^ and two used a Social Return on Investment (SROI) approach,^2-3^ which has a broader (e.g. social, economic and environmental) concept of resulting value (Nicholls, Lawlor et al. 2012). Six of the included studies included a review of the effectiveness of the implementation of the co-located welfare advice services.^2,6-7,9-10,12^ Nearly half of the reviewed studies explored participant experience of the intervention. Recipients of welfare advice were most commonly studied (n=7),^1-2,4-5,9,10,14^ alongside healthcare professionals working in the setting (n=6).^1-2,5,8-9,14^ Two studies examined the experiences of welfare advisors delivering the intervention.^1-2^

The quality of over half of the papers was assessed as high (n=5) ^1,10-12,14^ or medium (n=3).^4,6,8^ These better quality studies used robust approaches and made attempts to adjust for observed confounders. The quality of the remaining six studies was assessed as low, owing to a lack of reporting of their methodological and analytical approaches.^2-3,5,7,9,13^ The majority of reviewed studies were assessed as being of high relevance to the review objectives^2-12,14^ (n=12), with two studies being assessed with medium relevance.^1,13^ Half of the included studies were assessed as thick on the ‘richness’ of their findings.^1,4,6,10-12,14^ Studies of high or medium quality were also usually found to be thick on the assessment of the ‘richness’ of their findings. No studies were rejected on the basis of their quality, relevance or richness of their findings.

### Study findings

#### Baseline characteristics

The baseline characteristics of participants were similar across the four studies where they were reported (Table 4). They were more likely to be female, with an average age of 46 years. Few individuals under the age of 24 years sought access to welfare services. Details regarding the ethnicity of participants were reported in limited detail across four of the included studies; the majority of participants accessing welfare services described their ethnicity as white (74%).

**Table 4.** Baseline characteristics of participants across included studies

|  | Average | Number of studies |
| --- | --- | --- |
| **GENDER** |  |  |
| Male | 44% | 4<sup>1,10-11,13</sup> |
| Female | 56% | 4<sup>1,10-11,13</sup> |
| **AGE** |  |  |
| 17-24 years | 9% | 4<sup>8-9,11,13</sup> |
| 25-34 years | 11% | 4<sup>8-9,11,13</sup> |
| 35-44 years | 16% | 4<sup>8-9,11,13</sup> |
| 45-54 years | 22% | 4<sup>8-9,11,13</sup> |
| 55-64 years | 21% | 4<sup>8-9,11,13</sup> |
| 65+ years | 18% | 4<sup>8-9,11,13</sup> |
| Mean age | 46 years | 4<sup>8-9,11,13</sup> |
| **ETHNICITY** |  |  |
| BME communities | 20% | 4<sup>6-7,11,13</sup> |
| White | 74% | 4<sup>6-7,10-11</sup> |
| Not-specified | 6% | 4<sup>6-7,10-11</sup> |
| **HOUSEHOLD INCOME** |  |  |
| Income (<£4800 per annum*) | 51% | 2<sup>11,13</sup> |
| Income (£4800-£12000 per annum*) | 37% | 2<sup>11,13</sup> |
| Income (>£12,000 per annum) | 12% | 2<sup>11,13</sup> |
| **RELATIONSHIP STATUS** |  |  |
| Co-habiting | 38% | 3<sup>1,10,13</sup> |
| Single | 51% | 5<sup>1,10-11,13</sup> |
| Other | 11% | 2<sup>1,10</sup> |
| **EMPLOYMENT STATUS** |  |  |
| Employed | 19% | 2<sup>1,11</sup> |
| Not working due to long term illness or disability | 42% | 2<sup>4,10</sup> |
| Looking after the home | 3% | 2<sup>1,10</sup> |
| Unemployed | 18% | 4<sup>1,4,10-11</sup> |
| Retired | 18% | 4<sup>1,4,10-11</sup> |

\* The threshold for claiming universal credit for single people over the age of 25 years old is £4800. The threshold for joint claimants of universal credit for people over the age of 25 years old is £4800-12000. Correct as of 22 August 2021.

#### FINANCIAL IMPACTS

The theory of change model (Figure 1) proposes that access to co-located welfare advice services and improved welfare leads to greater financial stability, through improved income, support with debt relief and greater financial literacy and an awareness of welfare rights. This component was supported by the studies included in this review. All studies included in this review highlighted that there were improvements in financial outcomes for individuals who access co-located welfare advice services. This was reported by participants, healthcare professionals and welfare advisors alike. Improved and greater stability of household income came from backdated payments from unclaimed benefits and regular gains in household monthly income, through successful applications for eligible benefits.^3,5-7,9-11,13-14^ Many participants also reported receiving debt advice or support in reducing their levels of debt following access to welfare services in the included studies.^6,9-10,13-14^

Several studies reported that participants felt that their knowledge about financial issues, the law and their rights had improved as a result of having access to a welfare advisor.^1,3-4,6-7,9,10,13^ They felt better able to deal with current and potential future welfare problems. Even participants who only received advice but did not gain financially reported feeling that their confidence in managing finances had increased. Studies report that those who accessed welfare services were also more likely to know where and how to access advice in the future, should they need it.^11-12^ They also reported knowing how to avoid financial support-seeking behaviours that are detrimental to financial security, such as using credit cards and overdrafts.

#### HEALTH AND SOCIAL IMPACTS

The theory of change model proposes that a welfare advice service co-located in a health setting improves health and wellbeing through three mechanisms: reduced mental health and stress-related conditions; reduced levels of non-communicable disease; and less communicable disease. Improved physical health, or the perception of such, was reported as a positive outcome in most studies included in this review by participants, healthcare professionals and welfare advisors alike.^1-3,5-6,8-14^

*“Most respondents [medical professionals and welfare advisors] acknowledged that where underlying social drivers affected patients’ health, health improvement would be unlikely through medical intervention alone*.*”*

Study 12

Several studies reported that participants and welfare advisors felt that access to co-located welfare services led to improvements in mental health and overall feelings of wellbeing, thus achieving a greater quality of life.^1-3,5-14^

For most included studies, impacts on mental health were explored using qualitative methodology, with two studies conducting a robust qualitative analysis using a thematic analysis^12,14^ and one using frequency counts of commonly reported outcomes.^4^ Two studies measured mental health and wellbeing outcomes using validated tools, comparing self-reported changes to mental health between an intervention and control group.^10-11^ These studies demonstrated improvements to mental health and wellbeing outcomes following intervention compared to controls. One study^10^ presented descriptive statistics owing to lack of statistical power and the second study^11^ presented outcomes as odds ratios, finding that mental health and wellbeing outcomes only improved significantly for recipients who were female or belonged to black ethnic groups. A meta-analysis for mental health and wellbeing outcomes was not possible due to heterogeneity in outcome measures utilised.

Where reported in included studies, improved mental health and wellbeing were attributed largely to reductions in levels of stress, by way of: improved income; ^3,5-7,9-11,13-14^ debt relief;^6,9-10,13-14^ and support with managing bills and finances.^6-7,9^

*“[CAB] was invaluable. I’d have killed somebody, or killed myself if I hadn’t got it sorted out because it was just going downhill*.*”*

Study 14

Three studies of varying quality assessment (low medium and high respectively) found that many of their participants reported a feeling of self-worth and security following use of the services.^4,6,11^ Two studies of medium and high quality assessment, found that there were fewer accounts of suicidal ideations and reduced need for medication as a result of improved mental health.^5,8^ One high quality study found statistically significant reductions in prescriptions for anxiolytics and hypnotics (42% reduction (P=0.016)) during the six months after referral to the service compared with the six months before and a non-significant reduction in nurse appointments (from 1.50 to 1.34 per participant), suggestive of improved mental health outcomes for participants accessing co-located services.^8^ However, this study found no change in appointments or referrals for mental health conditions. Where measured objectively, through access to GP consultation records and as a self-reported measure, there was a 27% average (range 22-31%) reduction in antidepressant prescribing following receipt of co-located welfare advice.^5,8^ One medium quality study^5^ used simple frequency counts of self-reported outcomes to collect this data and a second before and after study^8^ accessed GP records to measure frequency of GP consultations in the six months before and after intervention. Both studies presented their results descriptively owing to a lack of statistical power.

Further improvements in mental health were demonstrated by two studies, of medium and high quality, that reported that some participants felt that they were able to talk to family and friends after receiving welfare rights advice and this had improved close relationships.^6,14^ There were fewer arguments in the household and significantly less stress within relationships. One low quality study found there was evidence to suggest that access to welfare rights advice helped to remove some participants from situations where they were living with abusive partners.^13^ This was not described in significant detail but involved re-housing participants away from their abusive relationships and securing their financial situation.

One high quality study included in the review demonstrated that participants who accessed welfare services also reported reduced substance misuse.^12^ This was facilitated by an improved access to primary care, mental health and community drug and alcohol services. Where housing conditions were poor, some participants reported reduced drug and alcohol use following access to the welfare service through improved housing conditions and thus breaking the cycle of the resumption of alcohol and substance misuse.^12^

Two high quality studies highlighted that some general practitioners were more sceptical about the long-term improvements to mental health owing to an improved financial situation.^8,12^ They felt that the issue of poor mental health and financial insecurity and instability were multi-factorial, each contributing to the other, and solving the issue of poor mental health with a short term improvement in financial security would not be sufficient to solve the problem. This was also reported by some participants who still felt that they had significant money worries to contend with or who were still worried about the future.^8,12^

Several studies attributed improvements to physical health from addressing other social determinants of health.^1-3,5-6,8-9,11-14^ For all included studies, impacts on physical health was explored using qualitative methodology, with two studies conducting a robust qualitative analysis using a thematic analysis.^12,14^ No studies measured physical health using validated tools. Three high quality studies found that access to co-located welfare rights advice improved engagement with other community health services and thus improved compliances with treatment plans, particularly for chronic, complex disease management.^1,9,14^ Two studies, of medium and high quality, found that participants reported overall improved levels of nutrition and greater food security through improved income and access to alternative food sources, such as food banks.^6,14^ Several studies reported improved housing conditions for participants through assistance with housing applications and grants by welfare advisors.^1,3,6,9,11,13^

#### HEALTH SERVICE BENEFITS

Finally, the theory of change model also suggests that access to co-located welfare rights advice and improved welfare provides benefits to the NHS, through reduced primary and secondary care caseload, resulting in cost savings for the NHS and freeing up the resources required for those most in need.

Many studies, utilising qualitative methodology, reported that GPs and other administrative staff found co-located services to be time saving for doctors and administrative staff alike. Services reduced practice staff time spent on non-health issues both inside and outside of consultations, where this linked to direct rather than indirect support, such as reducing bureaucratic pressure involved with form-filling, rather than addressing problems such as depression linked to debt. ^2-3,5-8,11-12,14^

However, the studies included in this review suggested that there was a mixed experience of whether co-located welfare advice services reduced contact time with healthcare professionals. These studies were limited to a primary care setting. Where explored qualitatively, two studies, of medium and high quality, found that patients reported a reduced need for repeat GP appointments following access to co-located welfare rights advice.^5,12^ In two high quality qualitative studies, they found that there was a difference in experience of the services and its perceived effect on consultation rate by GPs.^12,14^ Some GP’s felt that the service had no impact upon their consultation frequency and in fact felt that it was their role to consider and to support patients with their social problems where they impacted upon health, despite others stating this was outside their clinical role and feeling unqualified to address them directly.^12^ Some participants reported booking additional GP appointments, where they might not have done otherwise, because they were in the building seeing the welfare advisor.^14^ Others report perceiving the welfare service as ‘an extra’ rather than instead of consulting their GP.^12,14^

Where measured objectively, through access to GP consultation records and as a self-reported measure, there was a 7% average (range 0-13%) reduction in GP attendance following receipt of co-located welfare advice.^5,8,11^ One high quality paper demonstrated a statistically significant reduction in GP attendance (13.1% reduction, P=0.017) for advice recipients in the six months after being in receipt of the intervention, compared to the six months prior, using a before and after study design.^8^ However one high quality paper found no difference in GP consultation rate in the three months following receipt of the intervention compared to a control group, using a quasi-experimental study design. One high quality paper using a before and after design found there was no difference in referrals to mental health services in a six month period before and after benefitting from co-located welfare rights advice.^8^

Several studies found that there was a high sense of achievement reported by healthcare professionals who engaged with co-located with welfare rights advice services.^2,6-8,12^ In one medium and two high quality studies, many reported a frustration with their inability to support patients with wider determinants of health and being able to refer into a service providing this support gave the health professionals a feeling of satisfaction. ^7-8,12^ Two low and one medium study reported that healthcare professionals referring into the service felt that their own financial literacy had improved as a result of their interaction with the co-located service, though there was no description of how this idea was explored with these healthcare professionals.^2,6,13^

#### CO-LOCATED SERVICES AS A SPECIALIST SERVICE

The theory of change model suggest that there are several mechanisms through which welfare advice services co-located in a health setting operate to increase uptake of advice and ultimately improve welfare, compared to welfare advice services offered in a conventional setting, owing to the nature of its co-location. This element was not a specific research question explored by the studies included in this review. However, through qualitative exploration of the impact of co-located services on participants, healthcare professionals and welfare advisors alike, a number of findings emerged that contribute to this theory.

Some of the included studies found that welfare advisors involved in the provision of co-located services felt that co-located services gave a greater sense of confidentiality and trust to participants, which was reflected by the views of participants in these studies.^2,6-7,9,10,12-14^ The authors of some studies, including several high quality studies, reported that provision of welfare services co-located within a health setting were also more able to target and reach some of the most vulnerable people, in comparison to conventional services not co-located in a health setting.^1-2,6-9^ The authors identified that health services and healthcare professionals often have a unique access to vulnerable individuals and can strengthen the identification of need for advice among these groups, thereby mitigating poverty and reducing health inequalities.

#### ECONOMIC EVALUATION

Nine studies provided information on 14,468 participants who accessed and were supported by the welfare rights advice services.^1,3,5-7,9-11,13^ Some studies went on to provide further details on the costs of the service provided to commissioners and the financial gains for the participants, NHS and wider society (Table 5). ^1,3,5-7,9-11,13^

**Table 5.** Economic evaluation of co-located welfare services

|  | Total | Average | Median | Range | Number of papers |
| --- | --- | --- | --- | --- | --- |
| **SERVICE USE:** |  |  |  |  |  |
| Participants supported | 14468 | 1608 | 622 | 19-6785 | 9<sup>1,3,5-7,9-11,13</sup> |
| Number of contacts | 23070 | 7690 | 1231 | 28-21811 | 3<sup>1,3,10</sup> |
| Number of issues resolved | 30347 | 15174 | - | 1725-28622 | 2<sup>3,13</sup> |
| Average number of contacts per client | - | 4 | 3 | 1-8 | 4<sup>1,3,10,13</sup> |
| Average number of issues per client | - | 4 | - | 3-4 | 2<sup>3,13</sup> |
| **SERVICE COST:** |  |  |  |  |  |
| Cost of service (per annum) | - | £660,324 | £843,597 | £79,000-1,058,375 | 3<sup>3,6,11</sup> |
| Cost of service (per person) | - | £272 | £272 | £124-421 | 4<sup>3,6,11,13</sup> |
| **FINANCIAL GAINS:** |  |  |  |  |  |
| Participant financial gains (per person) | - | £1,840 | £1,394 | £776-3,656 | 6<sup>1,3,6-7,11,13</sup> |
| Average income increase (per annum) | - | £2,757 | £3,046 | £963-4,274 | 6<sup>1,3,6-7,11,13</sup> |
| Debt managed (per annum) | £4,653,309 | £2,326,655 | - | £129,000-4,524,309 | 2<sup>3,9</sup> |
| Social return on investment (per £1 spent) | - | £27 | - | £15-39.00 | 2<sup>2,11</sup> |

Participants in receipt of general welfare rights advice had on average four contacts^1,3,10,13^ and four issues resolved per participant.^3,13^ Where reported, the majority of participants accessing the services received support on more than one issue.^3,13^

The average cost of this service per study to commissioners was £660,324 per annum, ranging from £79,000 to £1,058,375 per study.^3,6,11^ The average cost per client was £272 (£124-421).^3,6,11,13^ More established services were found to cost less owing to less funding being required for set up costs and efficiency savings.^2,6^

Financial gains ranged from one-off payments, owing to unpaid or incorrectly allocated benefits, to improvements in annual household income, as a result of successful claims for entitled benefits. Participants gained £1,840 on average in one off payments and also benefitted from an average increase of £2,757 in household income per annum across studies.^1,3,6-7,11,13^

Two services provided across three of the studies generated on average £27 of social, economic and environmental return per £1 invested. Both studies reported a positive return on investment that ranged from £15 to £39 return on investment per £1 invested.^2,11^

### Service implementation

Many of the studies described some of the factors they considered to have facilitated and/or hindered the successful implementation of a welfare advice service in a health setting. Figure 4 provides an overview of these factors, which are summarised in Table 3. Co-production of the services, effective communication, collaboration and integration and simple referral pathways, were some of the more recurring themes identified.

**Figure 4.**
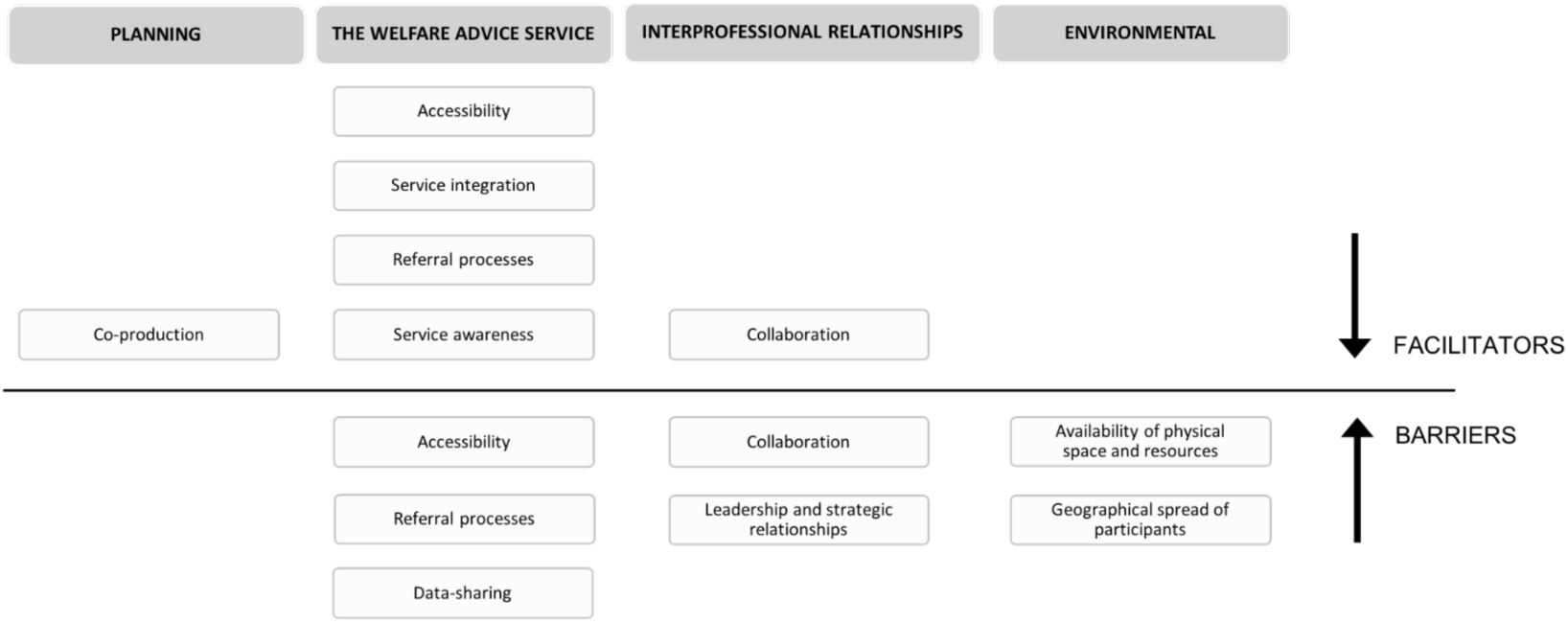
Factors affecting the successful co-location of a welfare advice service in a health setting

Co-production of the welfare advice service within the healthcare setting at the planning stages was seen as an essential factor for the successful implementation of the service.^6-7,13^ Involvement of both healthcare professionals and welfare advisors was found to be important, in order to raise awareness of the service amongst healthcare professions and thus improve appropriate referral rates.^6-7,13^ Several studies also reported the importance of higher level strategic buy-in to facilitate effective leadership and strategic working relationships.^6-7^ Co-production was felt to promote a more sustainable approach and built trust between the NHS and welfare services.

Most studies reported the importance of effective collaboration, communication and integration of the services.^2,6-7,9-10,12-13^ Some welfare advisors reported organisational barriers with NHS information sharing protocols which made referral processes more challenging and caused unnecessary delays.^6^ The quality of working relationships among project staff was also an important contributory factor in achieving successful implementation.^6,7,10,12^ Where working relationships were nurtured and created a welcoming, close and trusted relationship, the integrated services thrived. Welfare advice staff felt more integrated within the team when they shared physical space and resources with the healthcare staff, helping them to feel a part of the team.^8,12^

“*Co-ordination and collaboration do not happen on their own, that co-location is not just about the bricks and mortar. It is also about strategies to bring people together in a meaningful way*.*”*

Study 11

Simple referral pathways with clear associated documentation for professionals and participants improved referrals into the service. ^2,5,7,10,14^ The most common form of referral was directly by healthcare professionals, who are considered to know their patients well and are best able to identify need. ^2,5,7,10,14^ Referral by healthcare professionals legitimised the need for the services and helped to convey a sense of trust in the welfare service. ^2,10,14^ The option to self-refer was available in most services though it was not the most commonly accessed route.^12-14^

Finally, across many of the included studies, there was a strong sense that shared values (co-production, collaboration, communication, confidentiality, flexibility, holistic care and trust) between all involved with the services was important for a successful and effective service.. ^2,6-7,9,10,12-14^

## Discussion

### SUMMARY OF KEY FINDINGS

This systematic narrative synthesis review considers 14 research studies exploring the integration of welfare services within various health settings. The results of this systematic review are largely based on low level evidence of promise from qualitative and before and after studies, with only one study demonstrating causal evidence supporting the links between improved mental health and use of co-located welfare services.^11^ More research is needed using experimental methods and larger sample sizes.

Individuals accessing these services derived clear financial gains and reported improved financial security. Financial gains ranged from one-off payments, owing to unpaid or incorrectly allocated benefits, to improvements in annual household income, as a result successful claims for entitled benefits. Participants on average benefitted from an additional £1,840 in one-off payments and an additional £2,757 household income per annum. The services generated on average £27 of social, economic and environmental return on investment per £1 invested. This review highlighted that individuals accessing co-located services had improved knowledge about financial issues, the law and their welfare rights. This could suggest that co-located services are able to empower individuals, enabling them to better manage their finances and to improve future financial support seeking, rather than relying on overdrafts, credit cards and loans. This could in turn break the cycle of spiralling financial insecurity and ultimately reducing levels of poverty.

Improved health and wellbeing was reported as a positive outcome in most studies included in this review leading to a better quality of life. Where this was explored in greater detail, improvements to mental health and wellbeing were largely attributed to reduced levels of stress, owing to greater financial stability and security. This was achieved through improved income, debt relief and practical support with managing bills and finances. As a result of improved mental health, there was a reduced need for medication and fewer accounts of suicidal ideation. Individuals also felt that the quality of their relationships had improved owing to reduced levels of stress and many reported reduced substance misuse. Co-located welfare rights advice also improved engagement with other community health services and thus improved compliances with treatment plans, particularly for complex disease management, chronic diseases and the treatment of substance misuse. This systematic review also found evidence to suggest that co-located welfare rights advice reduces the workload for primary and secondary care services, resulting in cost savings for the NHS. This could suggest that co-located services are able to improve the availability of resources required for those most in need. Co-located services were found to support primary care in addressing key social determinants of health, something that was valued particularly by healthcare professionals. They were found to reduce the time spent by healthcare professionals on non-health issues, particularly by reducing bureaucratic pressure involved with form-filling. It was not clear whether co-located services reduced patient contact for healthcare professionals, with studies reporting mixed results.

This review contributes to the growing body of evidence that welfare rights advice in a health setting can improve health and wellbeing and benefit the NHS. The theoretical pathways through which the co-located service operates to benefit individuals are beginning to emerge. This review examined evidence to suggest how the nature of welfare rights advice co-located in a health setting might vary compared to conventional services and work to improve uptake of welfare advice and achieve these reported benefits to individuals. The findings from the review suggested that co-located services gave a greater sense of confidentiality and trust to participants. Provision of welfare services co-located within a health setting were also more able to target and reach some of the most vulnerable people, in comparison to conventional services not co-located in a health setting. The authors identified that health services and healthcare professionals often have a unique access to vulnerable individuals and can strengthen the identification of need for advice among these groups, thereby mitigating poverty and reducing health inequalities. These mechanisms were not explored as primary outcomes for the studies included in the review and many of the proposed mechanisms through which these services are proposed to work in the theory of change model have not yet been formally explored.

This review also contributed to our understanding on the implementation of welfare services in a health setting. Findings were consistent across the studies about the importance of co-production and collaboration from the inception through to the evaluation of the service. The review captured the importance of establishing an infrastructure for efficient and effective data sharing to support robust referrals processes that are visible and accessible to all individuals working within the organisation. Physical co-location of the service provided to participants was not sufficient for a successful and sustainable service. The need for effective integration of both the systems and the people working within both services was highlighted as equally important. Welfare advisors in particular needed to be provided with sufficient space and resources, comparable to other healthcare professionals, working within the organisation, in order to work effectively and feel part of the same team. Efforts to treat welfare advisors as equal to other healthcare professionals was also valued. A strong sense of values-based practice was demonstrated across the review and was associated with a successful welfare service. These were of co-production, collaboration, communication, confidentiality, flexibility, holistic care and trust.

The nature of the welfare advice service, how it operates within a health setting and how visible and accessible this service is to participants and professionals referring into the service were seen as important facilitators. The manner in which participants access the welfare services varied, from drop-in to referral by a health practitioner. Generally the preference amongst participants was for a timed appointment rather than a drop-in service, which was a perception shared by staff. Healthcare professionals and general practice staff report that patients are more likely to access services when advised to if they make a timed appointment at the point of referral, rather than relying on a drop-in service, as they seem to feel more accountable to the appointment. Being able to refer patients to a co-located welfare advice was seen as a valuable asset for patients and healthcare staff alike and sets co-located services apart from some other services. Many reports of the research reflected upon the challenges associated with evaluating a welfare service. This included challenges in data collection, with existing NHS information and data sharing protocols posing a particular challenge. Many struggled to recruit sufficient participants or were unable to follow-up sufficient numbers to enable a sufficiently powered study. Some implementation outcomes were particularly difficult to assess due to challenges in recruiting healthcare professionals into the studies. Many studies reported challenges in identifying suitable effectiveness and implementation outcome measures, resulting in significant heterogeneity in reported outcomes throughout the review. The challenge of recruiting minority groups into the study was also raised as a particular concern in many studies, despite efforts being made to address this.

## FINDINGS IN CONTEXT

This review updated and widened a 2006 review looking at the health, social and financial impacts of welfare rights advice delivered in healthcare settings (Adams, White et al. 2006). They found that there was evidence that welfare advice delivered in healthcare settings resulted in financial gains but there was limited evidence to suggest that this resulted in measurable health or social benefits. They also found little evidence of adequate robustness and quality to indicate that the impact goes beyond improving income.

Only one study has been published since that review with adequate power.^11^ It found that co-located welfare advice improves short-term mental health and wellbeing, reduces financial strain and generates considerable financial returns, compared to control groups. Qualitative methods have largely been favoured to explore the effects of co-located services on health and wellbeing.

Since the Coalition Government’s first Budget in 2010, significant reforms have been made to the UK’s social security system. Over the past decade, the nature and scope of benefits have changed significantly. For individuals this means that their benefits entitlement may have changed over time, in addition to the way in which they access them. The changing landscape of the social security system can generate confusion for those already accessing benefits, as well as those who may be entitled to them.

The COVID-19 pandemic has created or worsened financial difficulties and insecurity for the most vulnerable in society and further temporary and some more-permanent changes have been made in response to the pandemic and will continue to evolve over the course of the pandemic. During this time, access to financial support services and packages from the government has also been challenging for many families (Islam, Rahman et al. 2020). For the most vulnerable groups, such as refugees and asylum seekers, face-to-face access to organisations for support with welfare and housing has been curtailed, which is how these services would normally be accessed (Dickerson, Kelly et al. 2020).

## LIMITATIONS

This review includes a wide range of studies utilising a variety of methodological approaches, statistical techniques and outcome measures. A large proportion of the studies included in this review were grey literature, not published in peer reviewed journals. Quality assessment of these studies was challenging as the methodological approaches were not well described. Although most of these studies were found to be of limited scientific quality, it was felt that it was important to include these studies in the review, as they often included legitimate data on financial outcomes and population coverage of the services and ensured the review was representative of the available evidence base. However, as grey literature is not well indexed, it is also difficult to be sure that all available evidence has been accessed, despite the systematic approach to both the search strategy.

Across all of the included studies, there was significant heterogeneity in the research methodology and outcome measures utilised, preventing robust comparison of effect between studies. Where studies evaluated changes in mood, each study included in this review used a different measure of depression, levels of anxiety or measure of wellbeing. There is also a lack of statistical analyses of outcomes presented from service evaluations with the majority reporting simple descriptive measures. These variations and limitations meant that it was inappropriate to perform formal meta-analysis and the interpretations and conclusions drawn from the review are more subjective in nature.

Finally, this review is limited to the United Kingdom given that health and welfare systems are country specific with significant variation existing between countries. Therefore the results from this systematic review may not be generalisable to other countries and should be interpreted with care. However, some conclusions may be applicable internationally, such as how the co-located services are implemented and evaluated.

### Areas for future research

This systematic narrative synthesis builds upon the previous body of evidence provided by the systematic review published by Adams et al. (Adams, White et al. 2006) and provides an update on the body of evidence emerging in this area since the UK welfare reforms in 2010. It highlights that the quality of research carried out in this area has not progressed significantly since the last review and that investment is needed in future research.

Future research in this area needs to be well resourced, sufficiently powered with a robust comparator group to build upon the theoretical models proposed in this review by the existing but limited body of evidence. This review highlights the need for this research to utilise common health outcome measures that can enable comparisons to be made across the literature and for economic evaluations incorporating both a patient and a health services perspective. In order to draw firm conclusions about the links between the provision of welfare advice and improvements in health and wellbeing and reducing health inequalities, research needs sufficient resources to follow-up patients over the short, medium and long term.

This review demonstrates a largely homogenous participant group across the included studies, with a significant under-representation of ethnic minority groups, with evidence suggesting that these are some of the most in need. Further research needs to be conducted to ensure co-located services are best able to reach those most in need and to explore the health and social impacts of the services for these groups.

This review included a greater number of studies with welfare services co-located in a primary care setting, which is perhaps reflective of the more established relationship between welfare service providers and primary care providers. However, this may also reflect a lack of formal evaluations conducted in a secondary care setting and research should be planned to ensure it reflects the scope of available services.

## Conclusion

This review contributes to the growing body of evidence that welfare rights advice co-located in a health setting can improve health and wellbeing and provides cost savings to the NHS, freeing up resources for those most in need. This review also examines how the literature builds the evidence base to support the proposed theoretical pathways through which the co-located services operate to reduce health inequalities.

This review demonstrates that welfare advice services co-located in health services generate significant financial gains for participants and for the first time shows wider welfare benefits to participants, including access to housing, food, transport and employment. This contributes to the theory that these welfare services both directly and indirectly address social determinants of health, thereby improving health and wellbeing and reducing health inequalities. Given the high number of included studies of low scientific quality, the interpretations and conclusions drawn upon in this review are considered subjective. There remains a need for high quality research in this area to further build upon this theory and to measure the strength of these pathways over time. Further work is also needed on how deliver a service that best meet the needs of minority groups who are under-represented in existing research.

Although performing some sort of evaluation of welfare services is often a requirement of funding, additional resources to support such evaluations is limited. Those commissioning and implementing welfare services co-located in health settings should consider investing additional funds and securing the appropriate skills to conduct a robust evaluation of service implementation and effectiveness, guided by the findings of this review.

## Data Availability

All data produced in the present study are available upon reasonable request to the authors

## Appendix one example search strategy

Search strategy for Medline via Ovid using keywords

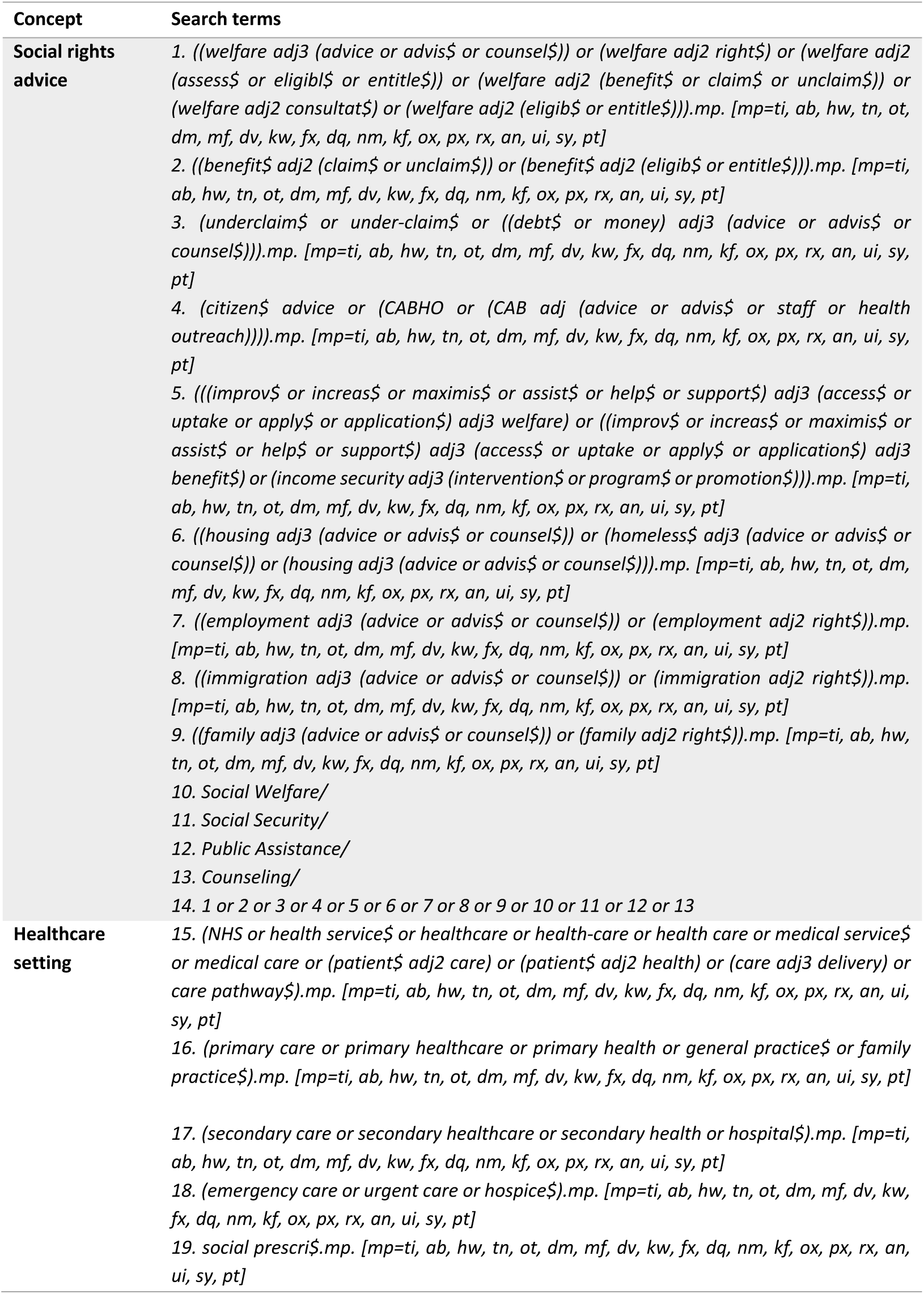
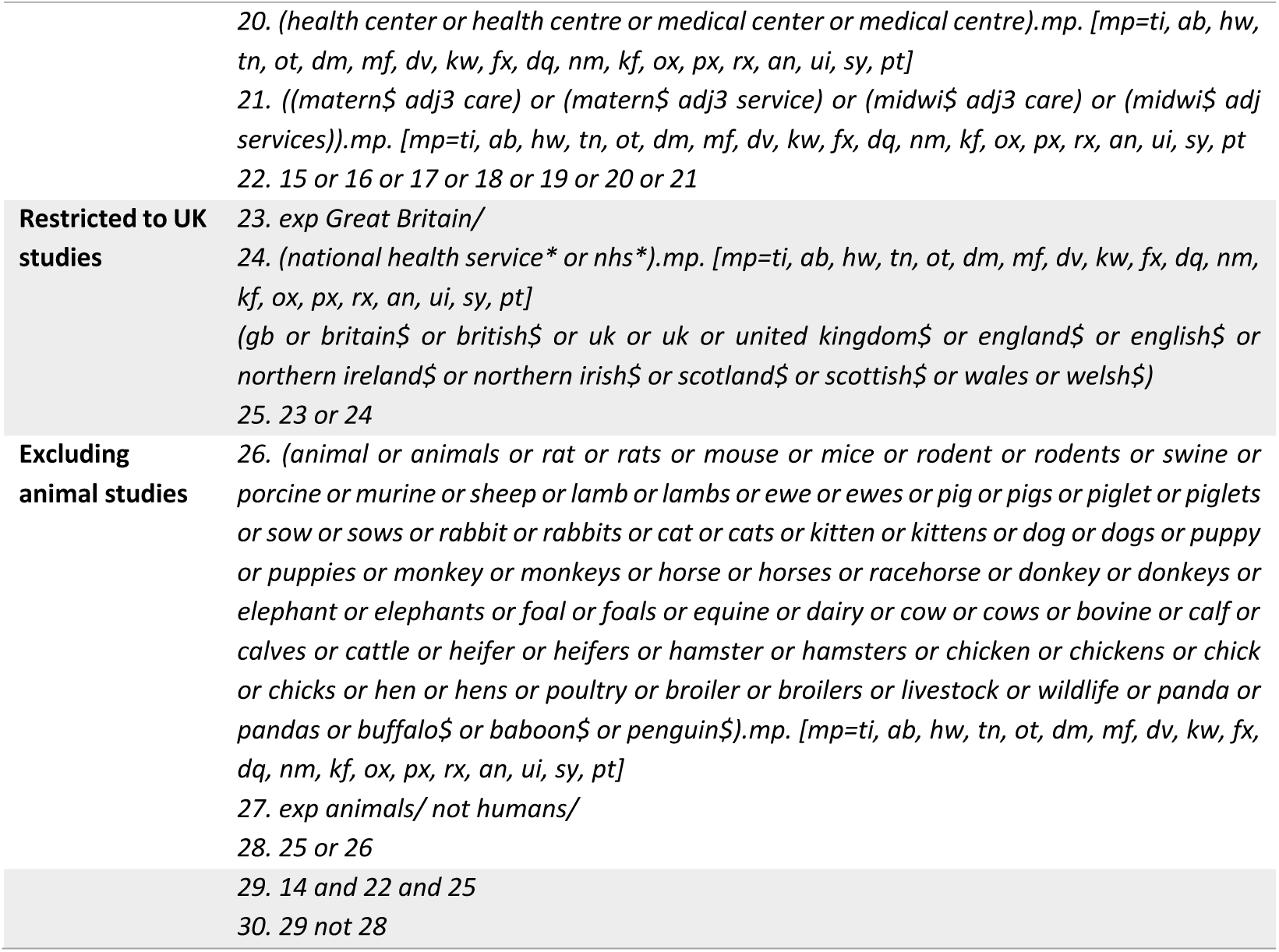

## Appendix two data extraction proforma

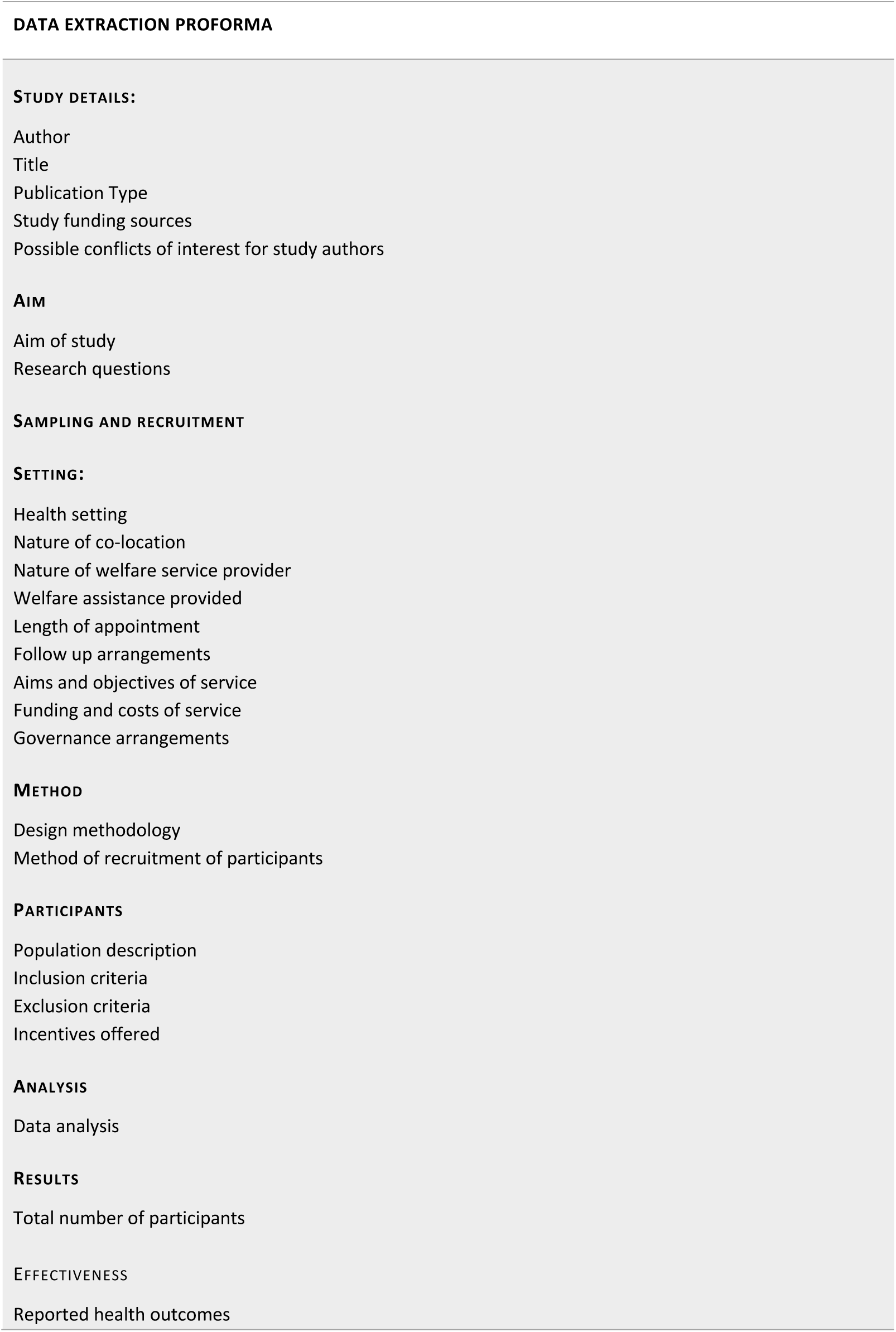
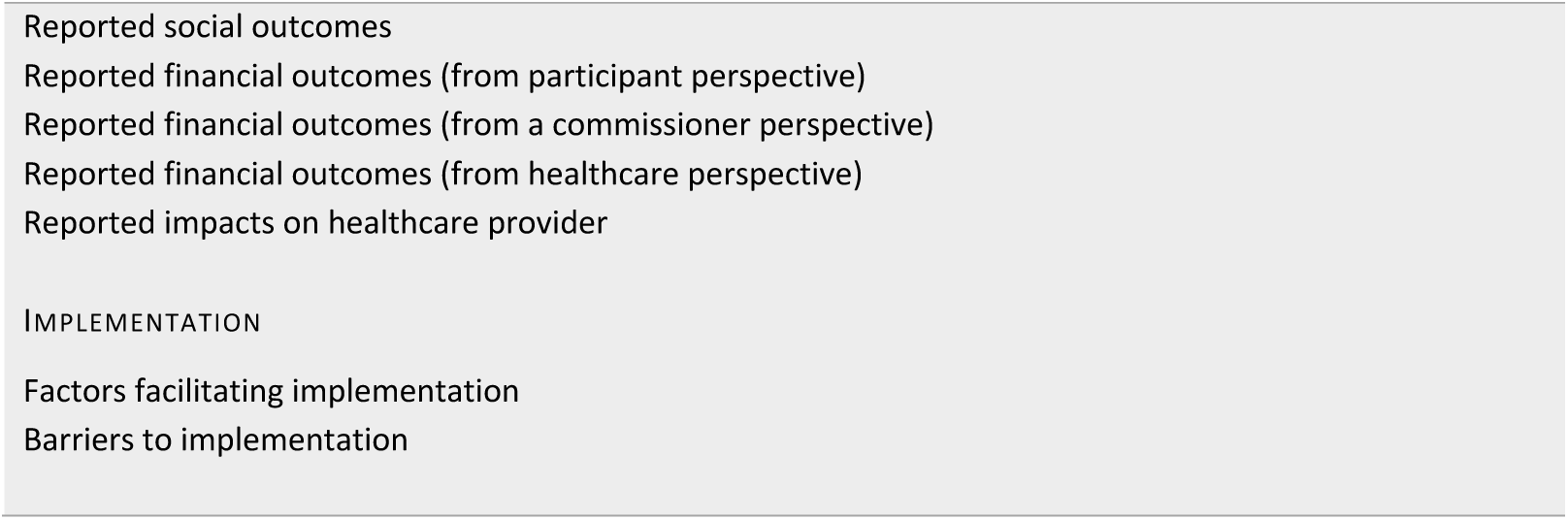

## APPENDIX THREE EXAMPLE OF A TEXTUAL SUMMARY

## Setting

The study was conducted in two urban primary care general practices in England.

## Intervention

The provision of co-located welfare rights advice services varied across locality. Co-located services in locality 1 provided specialist casework advice on welfare benefits and debt. They offered a walk-in “first-come-first-served” service that was open to all residents. In locality 2, the co-located welfare service offered booked appointments and casework advice on a broader range of issues e.g. housing and employment.

## Aim of study

To develop an initial programme theory for how the provision of co-located advice supports specific practice outcomes, and to identify salient barriers and enabling factors.

## Sampling and recruitment

GPs, practice managers, GP receptionists and advice staff from intervention practices in both localities invited to participate. Sampling aimed to include representatives from each job role as well as from both the advice and comparison groups. Further sampling also aimed to include a greater number of GPs.

## Study design

Twenty-four semi-structured interviews were conducted with general practice staff, advice staff and service funders between January and July 2016. This study is nested within a mixed-methods evaluation described elsewhere.^50^ Interviews were chosen rather than focus groups both due to practical difficulties of bringing together practitioners at the same time and to enable individuals in different roles within the same practices to speak freely.

## Data analysis

Data were thematically analysed and a modified Realist Evaluation approach informed the topic guide, thematic analysis and interpretation. The topic guide was built on a formative evaluation covering experiences, attitudes and expectations of the co-located advice service.

## RESULTS

Two outcomes are described linked to participant accounts of the impact of such non-health work on practices: reduction of GP consultations linked to non-health issues and reduced practice time spent on non-health issues. It was found that individual responses and actions influencing service awareness were key facilitators to each of the practice outcomes, including proactive engagement, communication, regular reminders and feedback between advice staff, practice managers and funders. Facilitating implementation factors were not limiting access to GP referral and offering booked appointments and advice on a broader range of issues responsive to local need. Key barriers included pre-existing sociocultural and organisational rules and norms largely outside of the control of service implementers, which maintained perceptions of the GP as the “go-to-location”.

## Notes

### Competing Interest Statement

There are no competing interests to declare.

### Funding Statement

SR is in receipt of the National Institute for Health Research Doctoral Research Fellowship. This work was supported by the UK Prevention Research Partnership (MR/S037527/1), which is funded by the British Heart Foundation, Cancer Research UK, Chief Scientist Office of the Scottish Government Health and Social Care Directorates, Engineering and Physical Sciences Research Council, Economic and Social Research Council, Health and Social Care Research and Development Division (Welsh Government), Medical Research Council, National Institute for Health Research, Natural Environment Research Council, Public Health Agency (Northern Ireland), The Health Foundation and Wellcome.

